# Identification of gene fusions associated with amyotrophic lateral sclerosis

**DOI:** 10.1101/2022.06.04.22275962

**Authors:** Yogindra Raghav, Allison A. Dilliott, Tiziana Petrozziello, Spencer E. Kim, James D. Berry, Merit E. Cudkowicz, Khashayar Vakili, NYGC ALS Consortium, Ernest Fraenkel, Sali M.K. Farhan, Ghazaleh Sadri-Vakili

## Abstract

Genetics is an import risk factor for amyotrophic lateral sclerosis (ALS), a devastating neurodegenerative disease affecting motor neurons. Recent findings demonstrate that, in addition to specific genetic mutations, structural variants caused by genetic instability can also play a causative role in ALS. Genomic instability can lead to deletions, duplications, insertions, inversions, and translocations in the genome, and these changes can sometimes lead to fusion of distinct genes into a single transcript. While such gene fusion events have been studied extensively in cancer, they have not been thoroughly investigated in ALS. We leveraged bulk RNA-Seq data from human post-mortem samples to determine whether fusion events occur in ALS. We report for the first time the presence of gene fusion events in several brain regions as well as in spinal cord samples in ALS. Although most gene fusions were intra-chromosomal events between neighboring genes and present in both ALS and control samples, there was a significant increase in the number of unique gene fusion in ALS compared to controls. Lastly, we have identified specific gene fusions with a significant burden in ALS, that were absent from both control samples and known cancer gene fusion databases. Collectively, our findings reveal an enrichment of gene fusion in ALS and suggest that these events may be an additional genetic cause linked to ALS pathogenesis.

## Introduction

Amyotrophic lateral sclerosis (ALS) is a lethal, adult-onset, neurodegenerative disease primarily affecting motor neurons in the motor cortex, brainstem, and spinal cord.^1, 2^ Genetics is an important risk factor for ALS, as 40-55% of familial ALS (fALS) are due to known genetic mutations,^3^ and over 50 causative or disease-modifying genes have been identified that are linked to disease, including but not limited to superoxide dismutase 1 (*SOD1*), TAR DNA binding protein (*TARDBP*), fused in sarcoma (*FUS*), and a hexanucleotide repeat expansion in *C9orf72*.^4, 5^ In addition, genetic risk factors also contribute to sporadic ALS (sALS); however, the causes of more than 80% of cases remains unknown.^4^ One major potential genetic cause of ALS may be structural variants, such as deletions, duplications, insertions, inversions, and translocations, which have not been systematically examined in ALS. A recent analysis of known ALS-causing genes demonstrated a role for structural variants in this subset of genes.^6^ Specifically, genomic structural variants in *C9orf72*, valosin-containing protein (*VCP*) and Erb-B4 receptor tyrosine kinase 4 (*ERBB4*) genes were shown to modify ALS risk, age, and site of onset as well as progression and survival, highlighting the role of structural variants in ALS pathogenesis.^6^ Similarly, repeat expansions, which are one type of structural variation, in the *C9orf72* gene as well as the medium CAG repeat in the *ataxin 2* (*ATXN2*) gene can cause ALS.^4^ Therefore, we hypothesized that a systematic, genome-wide analysis might reveal additional loci where structural variants contribute to the risk of ALS.

Recent studies have also demonstrated that genomic instability, mostly due to alterations in DNA damage repair (DRR), may be associated with ALS pathogenesis^7–10^ Of interest, DDR is now considered to be a unifying mechanism underlying neurodegenerative disorders^11^ and DNA damage is increased and accumulates in the aging brain.^12^ In ALS, dysfunction in the DDR mechanism, caused by endogenous sources such as reactive oxygen species^10^ or the inability for neurons to recognize or repair DNA damage^13–14^ can trigger onset or worsen disease progression. This has been demonstrated in animal models of ALS as well as by the accumulation of DNA damage in induced-pluripotent stem cell (iPSC)-derived motor neurons and ALS post-mortem brain and spinal cord samples.^5, 7, 14^ Importantly, ALS-associated genes such as *SOD1*, *TARDBP*, *FUS* and *C9orf72,* are involved in DDR. Specifically, SOD1 can alter DDR mechanism through regulation of transcription, while TARDBP and FUS maintain the balance between single- and double-strand break repair. Lastly, the G_4_C_2_ repeat expansion in *C9orf72* impairs ataxia-telangiectasia mutated (ATM) signaling^7^ which is critical for the activation of the DNA damage checkpoint during the cell cycle. Together, these findings demonstrate that alterations in genomic stability and DDR occur and may underlie ALS pathogenesis.

While genomic instability includes amplification, translocation, deletion, and inversion events in the genome,^13^ it can also result in gene fusions. Gene fusions are formed when two independent genes become juxtaposed due to structural rearrangements, such as translocations, deletions, and inversions.^15–16^ Historically, gene fusions are associated with cancers^17^ and cause pathogenesis by either gain or loss of function.^18^ Focusing on fusion events in cancer has significantly improved many aspects of clinical care, such as in their use as biomarkers to stratify patients, predict relapse, monitor disease post-treatment, and identify molecular subtypes of cancers.^19–20^ Importantly, fusion transcripts/proteins are also promising therapeutic targets.^21–22^ Here, we investigated the presence of fusion genes in ALS post-mortem central nervous system tissues using bulk RNA-Seq data sets.

## Methods

### Source of RNA-seq data

All RNA-Seq data used in this paper were previously generated by Target ALS and the New York Genome Center (NYGC) ALS Consortium and were shared with us under a collaborative research agreement. These data consist of RNA-Seq from the motor cortex (including medial, lateral, and unspecified), cervical spinal cord, thoracic spinal cord, lumbar spinal cord, frontal cortex, temporal cortex, occipital cortex, hippocampus, and cerebellum of ALS and control individuals. Information on the sample preparation, sequencing and quality control can be obtained from the Center for Genomics of Neurodegenerative Disease (CGND) at the NYGC. Importantly, quality control of the data accounted for high-fidelity base predictions, GC content, total read count, percent of duplicate reads, percent of rRNA, and potential sample contamination.

### Determining gene fusion events from bulk RNA-Seq

Gene fusion predictions were identified using STAR-Fusion v1.10.0 with default settings.^23^ STAR-Fusion uses the RNA-Seq read aligner, STAR,^24–26^ to align reads with command-line flags optimized for fusion detection. Briefly, chimeric reads from STAR alignment were isolated to begin fusion prediction. Chimeric reads occur when either (1) a portion of a read aligns to one gene and another portion of the same read aligns to a different gene (split) or when (2) each end of a paired read set aligns to different genes (spanning). Using these chimeric reads, STAR-Fusion uses all-vs-all blastn to remove false positive chimeric alignments that are caused by sequence similarity. Following all-vs-all blastn filtering, the remaining set of reads was considered for gene fusions. Candidate gene fusion pairs with only one split read or one spanning read pair were discarded. Using the Duplicated Genes Database, fusions involving genes that are likely paralogs of each other were also removed as these predictions may have been due to sequence similarity. If certain genes were found to have over 10 other genes as potential fusion partners, these genes were removed from consideration as being “promiscuous”. Recurrent fusions found in healthy RNA-Seq datasets, such as the Genotype-Tissue Expression project (GTEx), Illumina Human Body Map and 1000 Genomes RNA-Seq, were removed to limit the possibility of false positives. The full list of healthy RNA-Seq databases compared against can be found here: https://github.com/FusionAnnotator/CTAT_HumanFusionLib/wiki#red-herrings-fusion-pairs-that-may-not-be-relevant-to-cancer-and-potential-false-positives. Lastly, fusion candidates were filtered based on the number of reads providing evidence for the event. This was done using fusion fragments per million total RNA-Seq fragments (FFPM). Fusions with FFPM less than 0.1 (one evidence fragment per ten million total reads) were discarded as this ratio corresponds to the 99^th^ percentile of ratios identified for fusions in GTEx samples. Importantly, within each sample, a specific gene fusion can have multiple high-confidence breakpoints, which denote the base pair for each gene in the pair where the gene either ends or begins. To avoid counting fusions multiple times within the same sample in future analyses, the dataset was filtered to only include the most common breakpoint for each fusion in each sample. Hence, all downstream analyses were done with “breakpoint-unique gene fusions.” All gene fusion events were classified based on the regions involved, first, broadly into inter-chromosomal and intra-chromosomal fusions. The intra-chromosomal fusions were further classified into four subtypes: (1) local rearrangements, which were fusions where the genes are in an unexpected order given the strand of each gene in the pair; (2) not close proximity, which encompassed genes >100 kb apart; (3) neighbors, which were fusions that encompassed genes <100 kb apart and did not show evidence of gene orientation rearrangement; and (4) overlapping neighbors, which encompassed genes whose spans overlapped by at least one base pair.

### Dataset quality control

The dataset was filtered by ancestry to avoid any potential confounding factors. Specifically, bulk RNA-Seq samples from patients with greater than 80% European ancestry were kept in the final analysis cohort. Furthermore, principal component analysis (PCA) was used to determine if batch effects existed between samples based on a variety of co-variates, including project, sequencing platform, capture library preparation method, sample tissue of origin, subject ethnicity, and subject sex. The underlying matrix used for this analysis included all samples carrying unique fusion gene pairs found in our analysis cohort and the FFPM metric for that specific fusion and specific sample. Our analysis identified no batch effects when mixing data from both sources, indicating the datasets could be binned for downstream analyses (Supplementary Figure S1).

### Gene fusion enrichment analysis

The distribution of breakpoint-unique gene fusions carried per sample was compared between ALS and control samples using Welch’s t-test. Comparisons were performed both independent of sample tissue source and within each individual tissue source. Welch’s t-test was also used to examine the association between specific intra-chromosomal gene fusion subtypes and ALS across all tissues by comparing the number of breakpoint-unique gene fusions of each subtype carried per sample between ALS and control samples. Subsequently, Welch’s t-test was used to examine the association between specific intra-chromosomal fusion subtypes and ALS at a tissue-specific level. For all statistical analyses, no statistical comparisons were performed in the thoracic spinal cord, sensory cortex, or occipital cortex as there were too few (n < 10) tissue samples from controls.

Following initial enrichment analyses of the full gene fusion dataset, it was determined that multiple library preparation methods were used in the initial RNA sequencing. A portion of the samples were prepared using manual capture library preparation, meaning that a technician performed the library preparation by hand; whereas the remaining samples were prepared using an automated library preparation, which is performed by an automated robotic system, which is now the conventional approach. Although examination of the PCA did not demonstrate any significant batch effects from library preparation method; to be cautious, we subdivided the samples based on their library preparation method, and gene fusion enrichment analyses were repeated to ensure signals of enrichment were not technical artifacts driven by the methodology.

### Individual gene fusion burden analysis

We determined whether each pair of genes encompassed by a fusion, hereafter referred to as a “gene fusion pair”, found in our cohort was observed at a greater or lesser burden in ALS than control samples using Fisher’s exact test based on the counts of samples with or without the gene fusion pair. The test was first done using the sum of all counts observed across all tissues. The results were also filtered to include only the significant gene fusions absent from cancer fusion databases (https://github.com/FusionAnnotator/CTAT_HumanFusionLib/wiki#fusions-relevant-to-cancer-biology) and absent from the control samples in our cohort, hereafter referred to as “rare gene fusions.” Lastly, we performed burden analysis using Fisher’s exact testing on gene fusion pairs of each intra-chromosomal subtype that was significantly enriched in specific tissues. In this way were able to determine whether individual gene fusion pairs may be driving the signals of enrichment observed in the previous gene fusion enrichment analysis. The gene fusion pairs of each subtype that were identified in significant tissues were first binned, and a Fisher’s exact test was run for each intra-chromosomal subtype, followed by individual Fisher’s exact test on gene fusion pairs identified in each individual tissue for each intra-chromosomal subtype that demonstrated significant enrichment.

### Data visualization and statistical analysis

Statistical analyses were performed using R statistical software 4.1.1 (R Core Team, 2014) in RStudio 1.4.1717. Data visualization was performed using the *ggplot2* R package (v3.3.5).^27^ For all individual gene fusion pair burden analyses, corrected p-values were calculated using Bonferroni corrections based on the total number of breakpoint-unique fusions observed within the respective tissue(s) and significance was measured at an alpha-level of p < 0.05. Circos plots were generated by using the shinyCircos^28^ web interface (https://venyao.xyz/shinyCircos/). PCA was conducted with default flags in scikit-learn v1.0^29^ with Python 3.9.4 using the fit_transform() function and PCA biplots were rendered using matplotlib 3.4.2.^30^

### Study approval

The study was approved by the Partners Healthcare IRB. Written informed consent was obtained from all participants prior to study enrollment. Post-mortem consent was obtained from the appropriate representative (next of kin or health care proxy) prior to autopsy.

## Results

### Identification and classification of gene fusion events from RNA-Seq datasets

The RNA-Seq datasets from Target ALS and the ALS Consortium consisted of 367 individuals with ALS and 90 controls with several tissue samples collected per individual resulting in a total of 1,542 ALS and 249 control samples (Supplementary Table 1). Distribution of tissues across ALS and control samples is reported in Supplementary Figure 1. In total, 607 unique pairs of genes were observed to form fusions. There was a total of 21,872 breakpoint-unique gene fusions in ALS samples, and a total of 2,780 breakpoint-unique gene fusions in control samples (Figure 1). To ensure that there were no potential batch effects from project, sequencing platform, capture library preparation method, sample tissue of origin, subject ethnicity, and subject sex we performed a principal component analysis (PCA) the matrix of fusion fragments per million total RNA-Seq fragments (FFPM) values. The assessed co-variates introduced minimal variance in the gene fusion data (Supplementary Figures 2-7).

**Figure 1.**
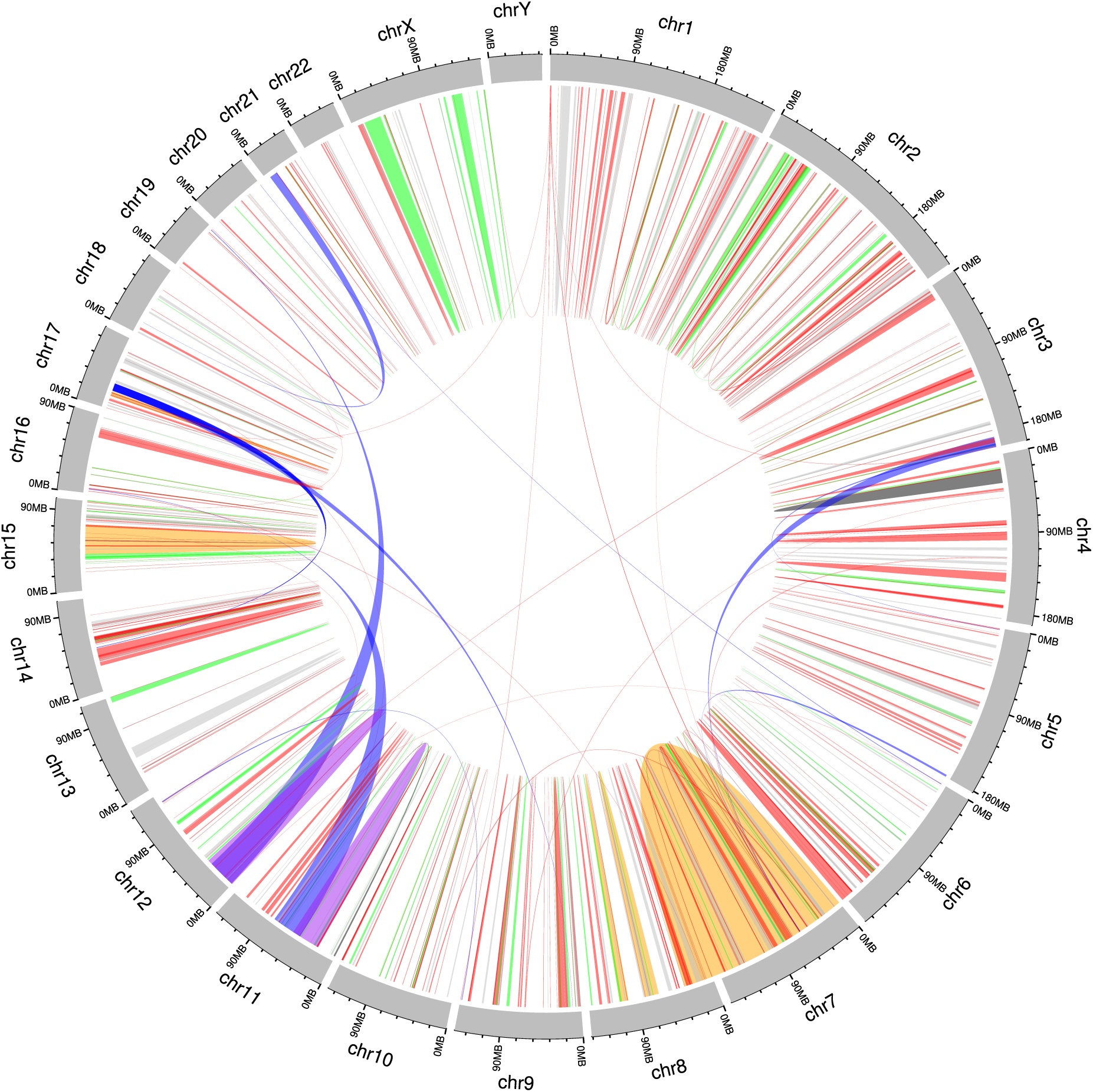
Circos plot of breakpoint-unique gene fusions identified in ALS and control samples using RNA-Seq datasets. Representation of significant fusion events where the width on the end of each line segment indicates the portion of the chromosome involved in the fusion event. Chromosomes were expanded 200X for clearer visualization and gene fusions on each are represented by color as follows: all tissues and per-tissue (black), significant in all tissues (orange), significant on a per-tissue basis (purple), inter-chromosomal (blue). Gene fusions are represented by lines on each chromosome that were expanded 10X for clearer visualization and include: ALS unique fusions (red), local rearrangements (brown), not close-proximity (green), and all others (gray).

To determine the origin of the gene fusions, we surveyed the proportion of inter-chromosomal versus intra-chromosomal events based on sample condition and found that most fusion events (>98%) were intra-chromosomal in both ALS and controls. We further divided these events into their intra-chromosomal fusion subtypes: local rearrangements, not close proximity fusions, neighbors, and overlapping neighbors (Figure 2A). Although most fusions were classified as neighbors in both ALS and control samples, we also identified a proportion of events classified as overlapping neighbor, not close proximity or local rearrangements in both ALS and control samples (Figure 2B). Furthermore, the chromosomes most often involved in the fusion events were chromosomes 6 and X, in both ALS and controls (Figure 2C-D). A summary of the different subtypes of gene fusions found per chromosome are displayed in Supplementary Figure 8.

**Figure 2.**
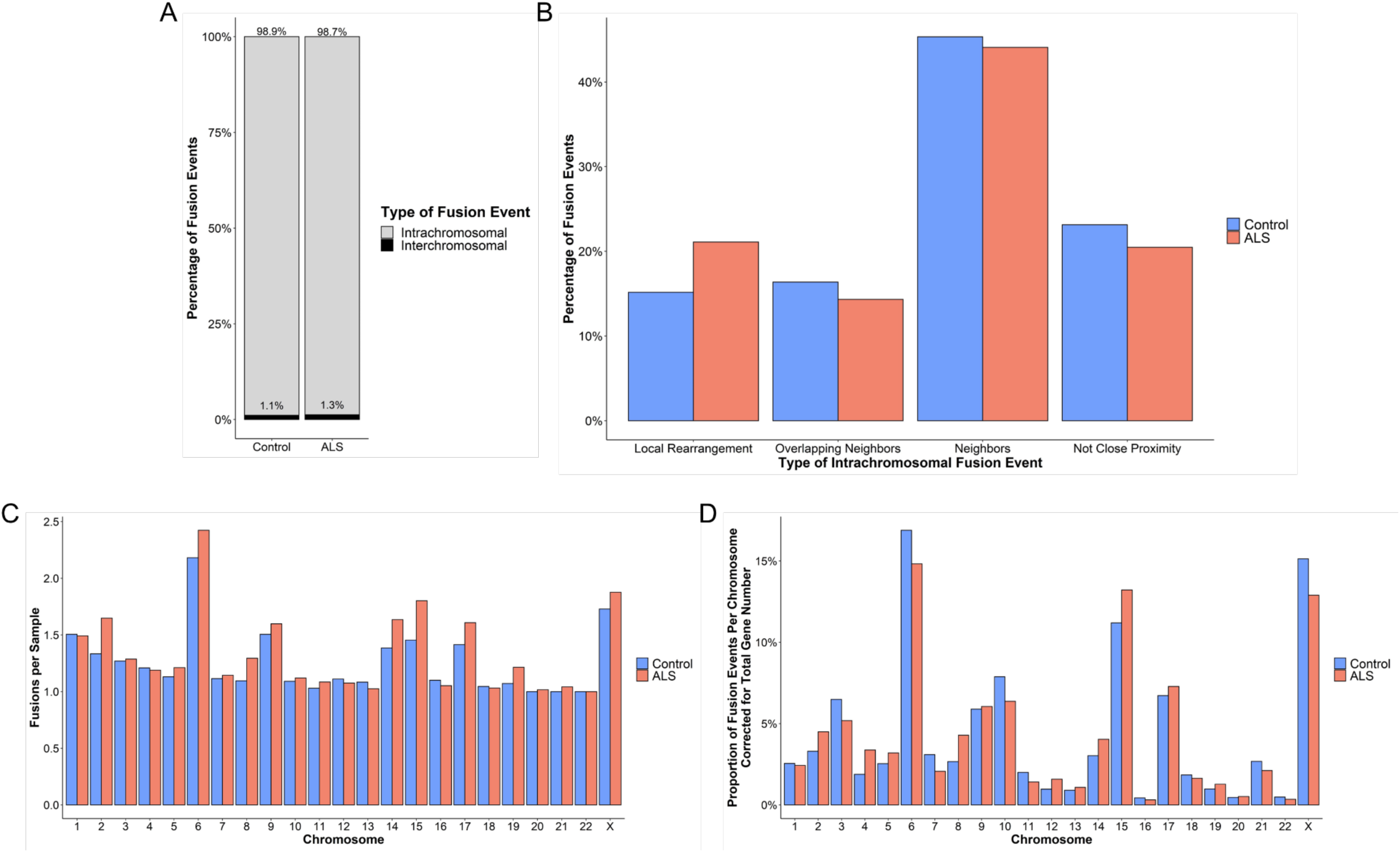
Characterization of the breakpoint-unique gene fusions and their subtypes in ALS and control samples (n = 1542 and n = 249, respectively). The breakpoint-unique gene fusions in ALS and control samples were compared to determine the distribution of (A) intra-chromosomal and inter-chromosomal gene fusions, (B) intra-chromosomal gene fusion subtypes, (C) fusion events per sample based on the chromosome(s) involved, and (D) the proportion of fusion events per chromosome(s) involved in the gene fusions corrected for total number of genes located on the chromosome (Ensembl, release 106).

### Distribution of gene fusion events between ALS and controls

To characterize the distribution of fusions, we compared the number of breakpoint-unique gene fusions carried by each ALS and control sample (Figure 3A). On average, ALS samples each carried significantly more breakpoint-unique gene fusion events than controls (mean ± SD: 14.18 ± 6.54 and 11.16 ± 6.10, respectively; Welch’s t-test, p = 4.505e-12). In particular, ALS samples each carried significantly more intra-chromosomal gene fusion events than the control samples (mean ± SD = 14.00 ± 6.45, and 11.04 ± 6.03, respectively; Welch’s t-test, p = 6.257e-12). However, there was no significant difference between the number of inter-chromosomal gene fusion events carried by each sample from ALS and controls (mean ± SD = 1.13 ± 0.36, and 1.07 ± 0.26, respectively; Welch’s t-test, p = 0.2677).

**Figure 3.**
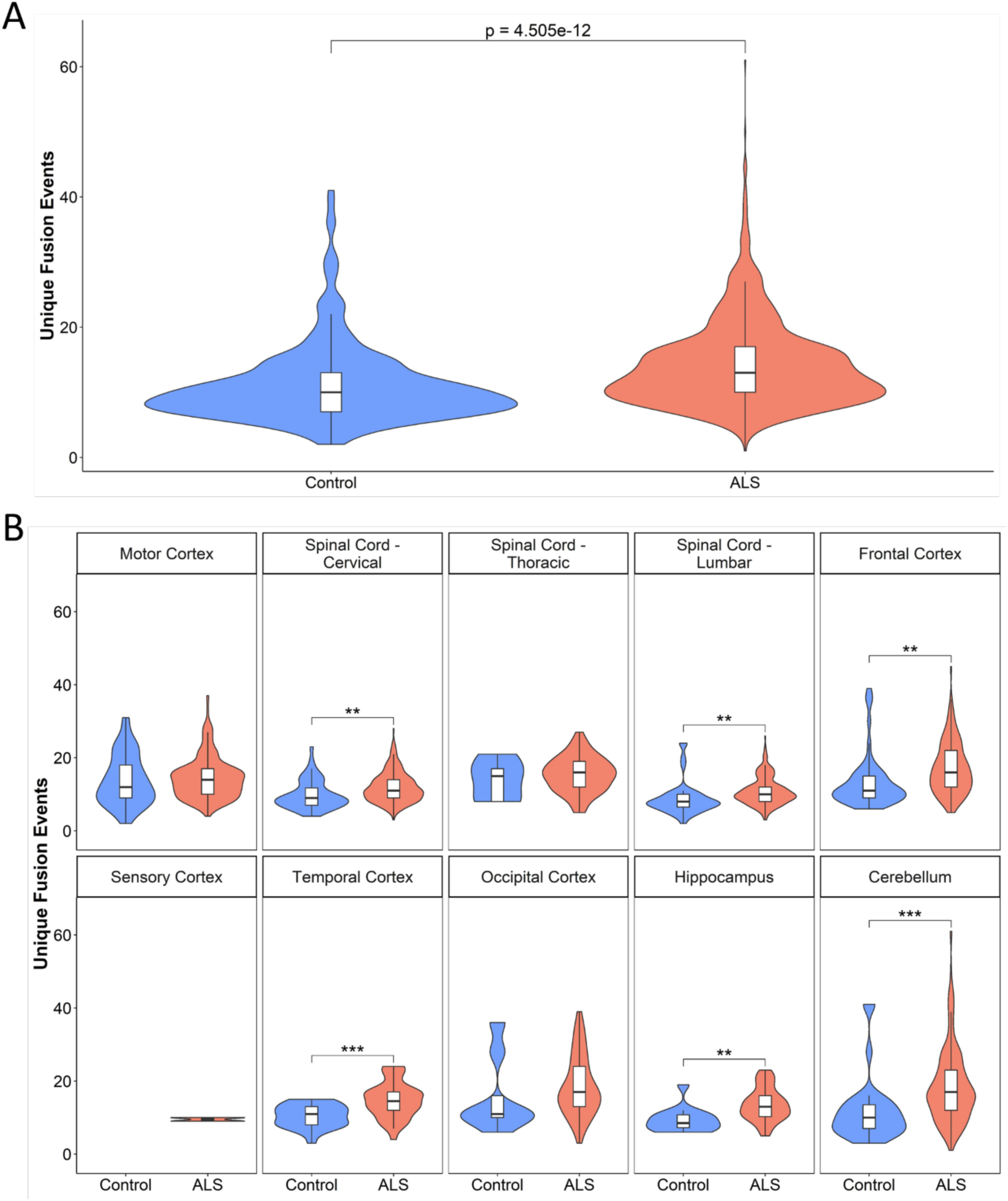
Breakpoint-unique gene fusions carried in ALS and control samples (n = 1542 and n = 249, respectively). The distribution of breakpoint-unique gene fusions carried per sample was compared between ALS and control samples using the Welch’s t-test, both independent of sample tissue source and within each individual tissue source. (A) ALS samples carried significantly more breakpoint-unique gene fusions (mean = 14.18; SD = 6.54) than controls (mean = 11.16; SD = 6.10) (p = 4.505e-12). (B) Significantly more breakpoint-unique gene fusions were carried by ALS samples compared to controls in the cervical spinal cord (p = 0.0022), lumbar spinal cord (p = 0.0012), frontal cortex (p = 0.0011), temporal cortex (p = 2.375e-4), hippocampus (p = 0.0056), and cerebellum (p = 1.393e-4). No statistical comparisons were performed in the thoracic spinal cord, sensory cortex, or occipital cortex as there were too few (n < 10) tissue samples from controls. * < 0.05; ** < 0.01; *** p < 0.001.

Next, we compared the number of breakpoint-unique gene fusions carried by each ALS and control sample within the individual tissue sources (Figure 3B). ALS samples had significantly more gene fusion events than controls as measured by Welch’s t-test in the following tissues: the cervical spinal cord (p = 0.0022), lumbar spinal cord (p = 0.0012), frontal cortex (p = 0.0011), temporal cortex (p = 2.375e-4), hippocampus (p = 0.0056), and cerebellum (p = 1.393e-4). There were no tissues in which control samples had more gene fusion events than ALS samples.

### Enrichment of intra-chromosomal gene fusion events

We tested whether ALS samples were enriched for specific subtypes of intra-chromosomal gene fusion events (Figure 4). Across samples from all tissues, we identified a significant enrichment of all four intra-chromosomal subtypes in the ALS samples compared to the controls as measured by Welch’s t-test: local rearrangements (p = 8.979e-15), not close proximity fusions (p = 0.0105), neighbor fusions (p = 2.930e-09), and overlapping neighbor fusions (p = 0.0151).

**Figure 4.**
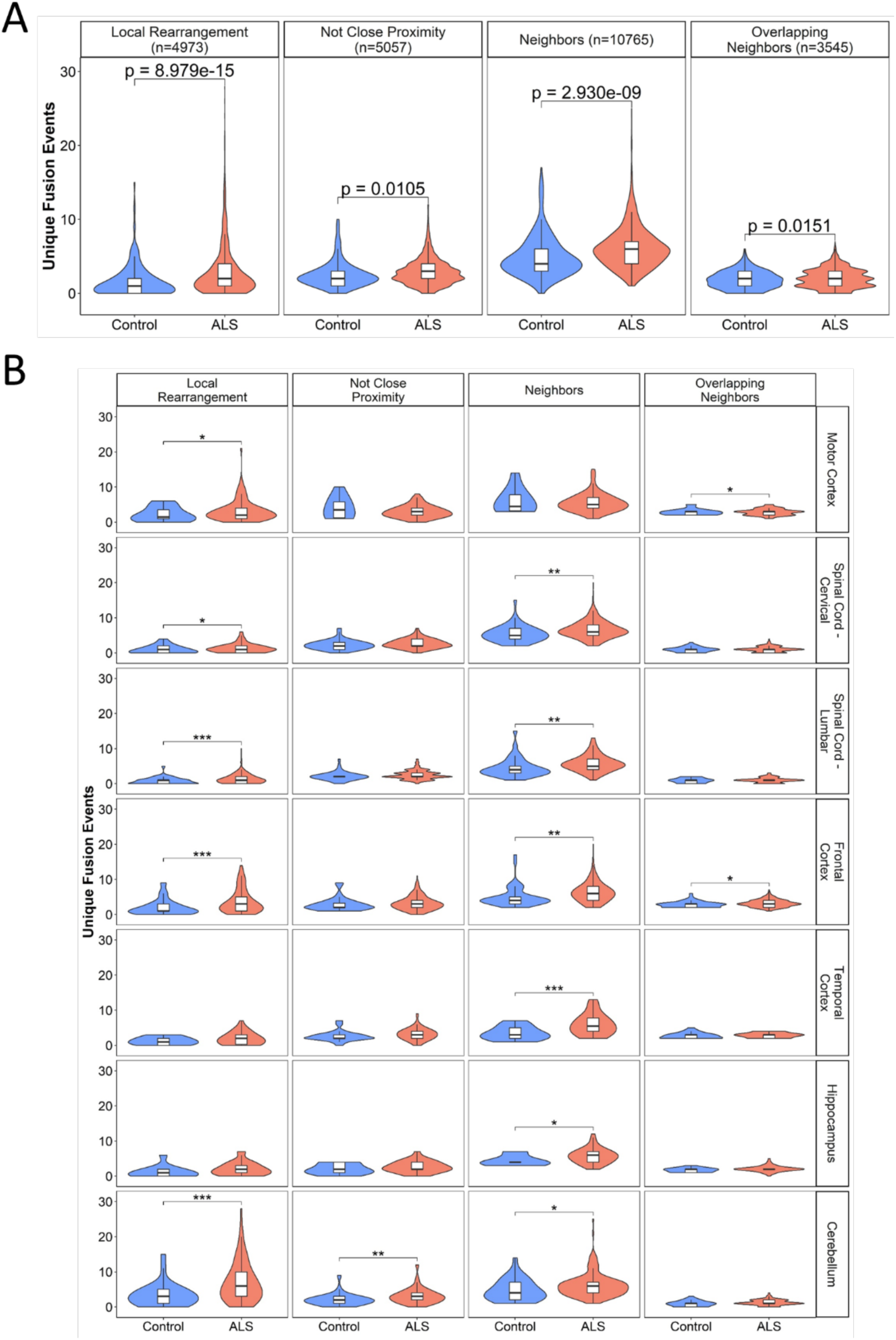
Enrichment of breakpoint-unique gene fusions carried by ALS and control samples. Welch’s t-test was used to compare the number of breakpoint-unique gene fusions of each subtype carried by each ALS and control sample both independent of sample tissue source and within each individual tissue source. (A) There was a significant enrichment of all four intra chromosomal subtypes in ALS compared to controls, including local rearrangements (p = 8.979e-15), not close proximity fusions (p = 0.0105), neighbor fusions (p = 2.930e-09), and overlapping neighbor fusions (p = 0.0151). (B) Following subgrouping of gene fusion based on the tissue source of the sample in which they were identified, a significant over-representation of local rearrangement events was identified in ALS samples from the motor cortex (p = 0.0206), cervical spinal cord (p = 0.0382), lumbar spinal cord (p = 8.997e-04), frontal cortex (p = 0.5.256e-05), and cerebellum (p = 5.367e-06). Not close proximity gene fusion events were significantly enriched in ALS samples from the cerebellum (p = 0.0055). Neighbor gene fusion events were significantly enriched in the ALS samples from the cervical spinal cord (p = 0.0021), lumbar spinal cord (p = 0.0080), frontal cortex (p = 0.0064), temporal cortex (p = 2.570e-04), hippocampus (p = 0.0155), and cerebellum (p = 0.0241). Finally, overlapping neighbor gene fusion events were significantly enriched in ALS samples from the motor cortex (p = 0.0458) and frontal cortex (p = 0.0222). No enrichment analyses were performed in the medial motor cortex, lateral motor cortex, thoracic spinal cord, sensory cortex, or occipital cortex as there were too few (n < 10) tissue samples from controls. * < 0.05; ** < 0.01; *** p < 0.001.

We carried out the same analysis separately for each tissue (Figure 4B). Local rearrangement events were significantly over-represented in ALS samples from the motor cortex (p = 0.0206), cervical spinal cord (p = 0.0382), lumbar spinal cord (p = 8.997e-04), frontal cortex (p = 5.256e-05), and cerebellum (p = 5.367e-06). Not close proximity gene fusion events were significantly enriched in ALS samples from the cerebellum (p = 0.0055). Neighbor gene fusion events were significantly enriched in the ALS samples from the cervical spinal cord (p = 0.0021), lumbar spinal cord (p = 0.0080), frontal cortex (p = 0.0064), temporal cortex (p = 2.570e-04), hippocampus (p = 0.0155), and cerebellum (p = 0.0241). Finally, overlapping neighbor gene fusion events were significantly enriched in ALS samples from the motor cortex (p = 0.0458) and frontal cortex (p = 0.0222).

We also repeated all of the above analyses separately for samples that had gone through automated library capture preparation (1047 ALS and 203 controls) and manual library capture preparation, which was a much smaller group (495 ALS and 46 controls) (Supplementary Figures 9-13). The results for the subset with automated library preparation largely matched the findings presented above. Our analysis demonstrated that in the manual subset there was a significant enrichment of local rearrangement gene fusion events in all ALS tissues compared to controls (Welch’s t-test, p = 0.0171), and the significant enrichment of local rearrangements in ALS cerebellum compared to controls (Welch’s t-test, p = 0.0381) that captured the findings from the automated sample set. The remaining discrepancies in the results were likely due to the much smaller sample size and lack of controls in the manually prepared samples.

### Individual gene fusion burden

Next, we aimed to identify whether individual gene fusion pairs were driving the enrichment of gene fusions in ALS samples compared to controls. We identified specific gene fusion pairs with a significantly greater burden of breakpoint-unique gene fusions in ALS or control samples by applying the Fisher’s exact test. Multiple testing corrected p-values were calculated using Bonferroni corrections based on the total number Fisher’s exact tests and the false discovery rate method.

To determine whether any individual gene fusion pairs were driving the general enrichment of fusions in ALS in comparison to controls, burden analysis was applied to the full dataset. The top ten results from the gene fusion pair burden testing performed across all tissue samples are presented in Table 1. Importantly, these top ten included the only gene fusion pairs that displayed a significant burden following Bonferroni correction when comparing ALS to control samples across all tissues. To highlight gene fusions that may be unique to ALS samples, we also filtered the gene fusion burden results to only include rare gene fusion pairs, defined as those absent from known cancer databases and from the control samples (Table 2).

**Table 1.** Gene fusion pairs with the highest individual burden in ALS versus control samples. Individual gene fusion burden tests were performed using the Fisher’s exact test. Bonferroni and FDR corrections were based on the total number of fusions across all tissues (n = 607). Abbreviations: ALS, amyotrophic lateral sclerosis; CI, confidence interval; FDR, false discovery rate; OR, odds ratio.

| **Fusion Name** | **Fusion Type** | **ALS**<br>**(n = 1542)** | **Control**<br>**(n = 249)** | **OR** | **Lower**<br>**95% CI** | **Upper**<br>**95% CI** | **p-value** | **Bonferroni**<br>**p-value** | **FDR**<br>**p-value** |
| --- | --- | --- | --- | --- | --- | --- | --- | --- | --- |
| AC006427.2--TAPT1-AS1 | Local Rearrangement | 471 | 34 | 2.78 | 1.89 | 4.19 | 8.76E-09 | 5.39E-06 | 3.48E-06 |
| KRTAP5-AS1--AP000867.5 | Not Close Proximity | 105 | 47 | 0.31 | 0.21 | 0.47 | 1.13E-08 | 6.95E-06 | 3.48E-06 |
| DOCK4--IMMP2L | Not Close Proximity | 0 | 8 | 0.01 | 0.00 | 0.09 | 1.27E-07 | 7.78E-05 | 2.59E-05 |
| AC090517.5--ZNF280D | Neighbors | 1116 | 140 | 2.04 | 1.53 | 2.71 | 5.02E-07 | 3.09E-04 | 7.72E-05 |
| TVP23C--CDRT4 | Neighbors | 940 | 110 | 1.97 | 1.49 | 2.61 | 7.70E-07 | 4.73E-04 | 9.47E-05 |
| AC009975.1--STON1-GTF2A1L | Local Rearrangement | 268 | 16 | 3.06 | 1.81 | 5.54 | 2.35E-06 | 1.44E-03 | 2.40E-04 |
| MAILR--ATP6V1C1 | Neighbors | 301 | 20 | 2.78 | 1.72 | 4.71 | 3.02E-06 | 1.86E-03 | 2.65E-04 |
| C8orf44--SGK3 | Neighbors | 202 | 11 | 3.26 | 1.75 | 6.74 | 2.06E-05 | 1.27E-02 | 1.59E-03 |
| PDC-AS1--ODR4 | Local Rearrangement | 222 | 15 | 2.62 | 1.52 | 4.85 | 1.18E-04 | 7.26E-02 | 8.07E-03 |
| FAM228A--FAM228B | Local Rearrangement | 59 | 0 | 20.01 | 1.21 | 318.39 | 1.85E-04 | 1.14E-01 | 1.14E-02 |

**Table 2.**
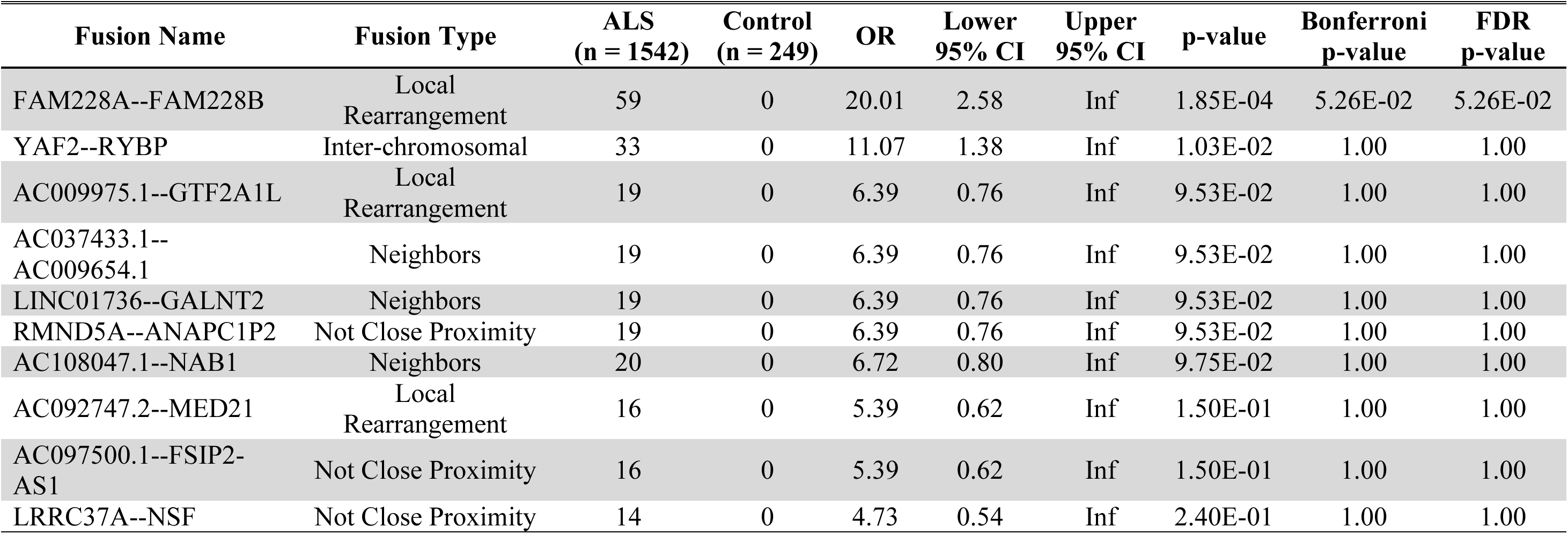
Rare gene fusion pairs with the highest individual burden in ALS versus control samples. Individual gene fusion burden tests were performed using the Fisher’s exact test and results were prioritized to identify only include rare breakpoint-unique gene fusions, which were defined as those absent from known fusion databases and absent from the control samples. Bonferroni and FDR corrections were based on the total number of rare fusions across all tissues (n = 280). Abbreviations: ALS, amyotrophic lateral sclerosis; CI, confidence interval; FDR, false discovery rate; OR, odds ratio.

Based on the results presented in Figure 4, we next aimed to determine whether specific gene fusions were driving enrichment of specific intra-chromosomal subtypes. To maximize statistical power and minimize potential signal from tissues not displaying enrichment of gene fusion pairs, we binned together all gene fusion pairs carried by samples from tissues displaying significant enrichments of fusion of that specific subtype. Therefore, samples from the following tissue sources were binned: motor cortex, cervical spinal cord, lumbar spinal cord, frontal cortex, and cerebellum in the burden test of local rearrangement fusions; cerebellum in the burden test of not close proximity fusions; cervical spinal cord, lumbar spinal cord, frontal cortex, temporal cortex, hippocampus, and cerebellum in the burden test of neighbor fusions; and motor cortex and frontal cortex in the burden test of overlapping neighbor fusions. We then performed an individual gene burden test for each intra-chromosomal gene fusion subtype using these binned groups of tissue sources (Table 3). Again, the gene fusion burden results were filtered to only include rare gene fusion pairs, defined as those absent from known cancer databases and from control samples (Table 4).

**Table 3.** Intra-chromosomal gene fusion pairs identified in tissues displaying significant enrichment in ALS samples with the highest individual burden in ALS versus controls. The individual burden tests of gene fusion events were only performed on fusions within tissues demonstrating significant differences in the number of gene fusions of each intra-chromosomal subtype carried by ALS and control patients. Only samples from tissues showing significant enrichment of that specific subtype were included in each burden test, including samples from the motor cortex, cervical spinal cord, lumbar spinal cord, frontal cortex, and cerebellum in the burden test of local rearrangement fusions; cerebellum in the burden test of not close proximity fusions; cervical spinal cord, lumbar spinal cord, frontal cortex, temporal cortex, hippocampus, and cerebellum in the burden test of neighbor fusions; and motor cortex and frontal cortex in the burden test of overlapping neighbor fusions. Bonferroni and FDR corrections were based on the total number of fusions observed within the respective tissues. Abbreviations: ALS, amyotrophic lateral sclerosis; CI, confidence interval; FDR, false discovery rate; n, total number of samples; OR, odds ratio.

| **Intra-chromosomal Fusion Type** | **Fusion Name** | **ALS (n)** | **Control (n)** | **OR** | **Lower 95% CI** | **Upper 95% CI** | **p-value** | **Bonferroni p-value** | **FDR p-value** |
| --- | --- | --- | --- | --- | --- | --- | --- | --- | --- |
| Local Rearrangement | AC006427.2--TAPT1-AS1 | 402 (1325) | 26 (200) | 2.91 | 1.89 | 4.66 | 7.95E-08 | 1.11E-05 | 1.11E-05 |
|  | AC009975.1--STON1-GTF2A1L | 234 (1325) | 16 (200) | 2.47 | 1.44 | 4.49 | 3.09E-04 | 4.33E-02 | 2.16E-02 |
|  | ADAMTSL3--SH3GL3 | 281 (1325) | 22 (200) | 2.18 | 1.36 | 3.63 | 5.72E-04 | 8.01E-02 | 2.53E-02 |
| Not Close Proximity | ABCD2--KIF21A | 67 (187) | 2 (29) | 7.49 | 1.79 | 67.06 | 1.12E-03 | 6.51E-02 | 9.79E-01 |
|  | PMS2P11--CCDC146 | 0 (187) | 2 (29) | 0.03 | 0.00 | 0.81 | 1.75E-02 | 1.00 | 9.79E-01 |
|  | AC023421.2--AC021517.2 | 0 (187) | 1 (29) | 0.05 | 0.00 | 6.05 | 1.34E-01 | 1.00 | 1.00 |
| Neighbor | TVP23C--CDRT4 | 633 (1047) | 82 (201) | 2.22 | 1.61 | 3.06 | 3.48E-07 | 6.74E-05 | 6.74E-05 |
|  | C8orf44--SGK3 | 159 (1047) | 8 (201) | 4.32 | 2.09 | 10.35 | 2.62E-06 | 5.08E-04 | 2.54E-04 |
|  | AC090517.5--ZNF280D | 728 (1047) | 109 (201) | 1.93 | 1.40 | 2.65 | 3.84E-05 | 7.44E-03 | 2.48E-03 |
| Overlapping Neighbor | AC127502.3--AC127502.1 | 91 (621) | 4 (85) | 3.47 | 1.26 | 13.37 | 1.02E-02 | 2.54E-01 | 2.54E-01 |
|  | AC067956.1--AC019211.1 | 131 (621) | 9 (85) | 2.26 | 1.09 | 5.26 | 2.05E-02 | 5.13E-01 | 2.56E-01 |
|  | CLDN12--CDK14 | 42 (621) | 3 (85) | 1.98 | 0.61 | 10.22 | 3.45E-01 | 1.00 | 1.00 |

**Table 4.**
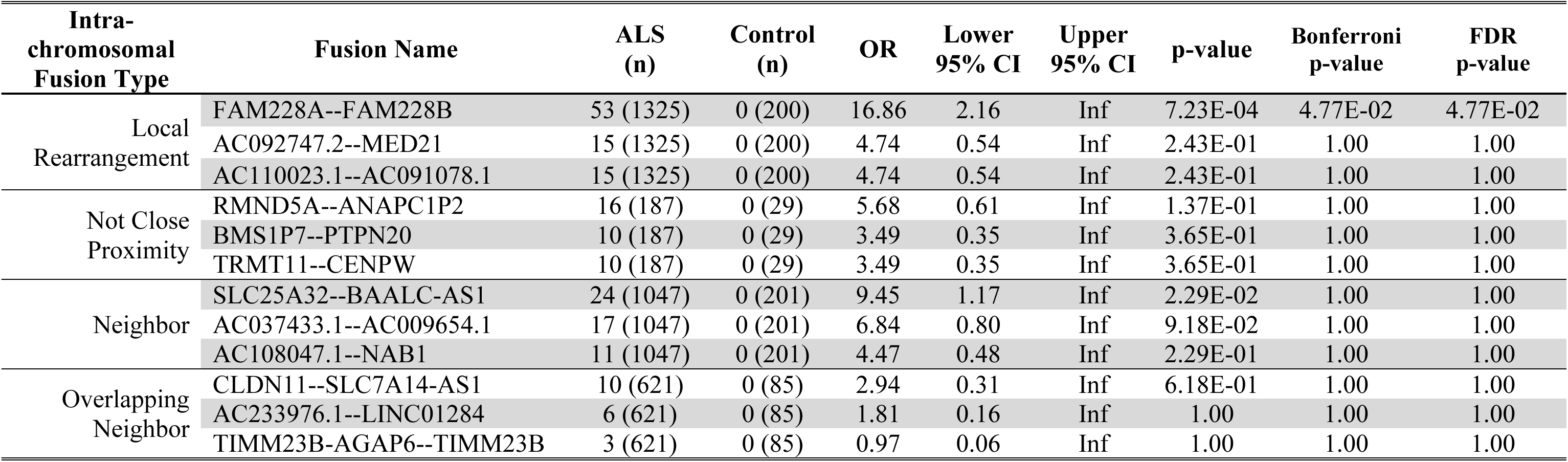
Rare intra-chromosomal gene fusion pairs identified in tissues displaying significant enrichment in ALS samples with the highest individual burden in ALS versus controls. The individual burden tests of gene fusion events were only performed on fusions within tissues demonstrating significant differences in the number of gene fusions of each intra-chromosomal subtype carried by ALS and control patients. Only samples from tissues showing significant enrichment of that specific subtype were included in each burden test, including samples from the motor cortex, cervical spinal cord, lumbar spinal cord, frontal cortex, and cerebellum in the burden test of local rearrangement fusions; cerebellum in the burden test of not close proximity fusions; cervical spinal cord, lumbar spinal cord, frontal cortex, temporal cortex, hippocampus, and cerebellum in the burden test of neighbor fusions; and motor cortex and frontal cortex in the burden test of overlapping neighbor fusions. Results were prioritized to only include rare breakpoint-unique gene fusions, which were defined as those absent from known fusion databases and absent from the control samples. Bonferroni and FDR corrections were based on the total number of fusions observed within the respective tissues. Abbreviations: ALS, amyotrophic lateral sclerosis; CI, confidence interval; FDR, false discovery rate; n, total number of samples; OR, odds ratio.

Finally, burden testing was performed for each intra-chromosomal subtype for gene fusion pairs identified in each tissue displaying significant enrichment of fusion of that specific subtype (Supplementary Table 2), to determine if individual gene fusions were driving the signals of enrichment in the tissue. Following multiple testing correction, significant burdens in ALS samples compared to controls were found for one local rearrangement in both the cervical spinal cord samples (*AC006427.2*--*TAPT1-AS1*; OR = 3.70 [1.62-9.55]; Fisher’s test, p = 0.0404, following multiple testing correction) and lumbar spinal cord samples (*AC006427.2*--*TAPT1-AS1*; OR = 5.96 [2.05-40.92]; Fisher’s test, p = 0.0075, following multiple testing correction), one neighbor fusion in the cervical spinal cord samples (*PAMR1--SLC1A2*; OR = 24.54 [3.00-Inf]; Fisher’s test, p = 0.0085, following multiple testing correction), one neighbor fusion in the temporal cortex samples (*AEBP2--AC024901.1*; OR = 32.02 [3.08-Inf]; Fisher’s test, p = 0.0112, following multiple testing correction), and one overlapping neighbor fusion in the frontal cortex samples (*AC067956.1--AC019211.1*; OR = 4.40 [1.52-17.48]; Fisher’s test, p = 0.0432, following multiple testing correction).

## Discussion

In this study, we leveraged RNA-Seq data from Target ALS and the NYGC ALS Consortium, and we report for the first time the presence of gene fusion events in ALS from several brain regions as well as spinal cord. Most fusions were intra-chromosomal events between neighboring genes and there was a significantly greater average number of breakpoint-unique gene fusion events identified per ALS sample compared to controls. Although fusion events were present in nearly all brain and spinal cord samples from both ALS and controls, they were significantly enriched in specific regions, such as cervical and lumbar spinal cord, frontal cortex, temporal cortex, hippocampus, and cerebellum in ALS compared to controls. Statistical comparisons could not be performed for the thoracic spinal cord, sensory cortex, or occipital cortex as there were too few tissue samples from controls. Lastly, we have highlighted specific gene fusions with a significant burden in ALS, including rare events that were absent from both known fusion cancer databases and from control samples in our cohort. Together, our findings demonstrate an enrichment of gene fusions that are unique to ALS, suggesting their potential involvement in the genetic etiology of the disease.

The overrepresentation of intra-chromosomal gene fusions in both ALS and control samples was consistent with trends that have been observed in several cancers, such as epithelial and prostate.^31–32^. Additionally, recent analysis of human cortex from healthy individuals revealed that most fusion events were formed from genes on the same chromosome.^33^ Although we also subtyped the intra-chromosomal gene fusions based on the proximity of the genes involved in the event, ALS samples were found to be significantly enriched for all four intra-chromosomal subtypes, including (1) local rearrangements, (2) not close proximity fusions, (3) neighbors, and (4) overlapping neighbors. We also identified specific gene fusion pairs within the tissues demonstrating significant enrichment of intra-chromosomal fusion subtypes, which require further investigation to determine their potential role in ALS. In some cases, these fusions demonstrated significant burden in ALS samples across all tissue samples, such as the local rearrangement *AC006427.2*--*TAPT1-AS1*, whereas other fusions demonstrated significant burden in ALS samples specifically in certain tissue types, such as the neighbor fusion in the cervical spinal cord samples *PAMR1-SLC1A2*. in the lumbar spinal cord, namely, which was found to have a significant burden in ALS samples (99/265) compared to controls (4/37).

Previously, gene fusions were largely detected using fluorescence *in situ* hybridization and quantitative real-time polymerase chain reaction; however, these methods do not allow for an agnostic screen of all potential fusion events. Rather, these methods specifically target known gene fusions.^34^ In contrast, RNA-Seq has proven to be an efficient method for detecting gene fusions across the entire transcriptome, yet technical limitations remain. One concern is the false positive associations that can result from RNA-Seq analysis. Recently, 23 RNA-Seq fusion detection methods were compared to examine accuracy as well as relative computational speed, and the STAR-Fusion algorithm was considered a top performer in both respects.^23^ Our choice of software was strategic to minimize the possibility of false positive findings. STAR-Fusion employs several filtration steps, including referencing against gene fusion databases from control populations to ignore fusions expected in healthy people. As the study of gene fusion events continues to grow, the establishment and extension of such databases remains imperative to gain a full understanding of the frequency with which the gene fusion events have been previously observed in respect to various phenotypes. For example, databases of expected fusions from brain tissues of non-neurological controls would have been particularly useful in this study. Lastly, we acknowledge the universal limitations of RNA-Seq, such as poor sensitivity for lowly expressed genomic regions and the delicacy of RNA samples that may affect sample quality and yield, of which the potential influence on gene fusion detection remains unclear.^35^

We have yet to determine if the newly identified gene fusions contribute to the development ALS as has been determined in oncology. Fusion genes are well-defined oncogenic drivers in several different types of cancer, demonstrating their potential for reprogramming of normal cellular function. Indeed, fusion events can lead to either gain or loss of function, causing overexpressed, constitutively active, or truncated products.^36^ ALS is approximately 50% heritable,^3^ yet the known ALS causing genes are present in less than 15% of patients. It is possible that some fusion events will explain the missing heritability. Many of the events that we identified are present at lower frequency in people without ALS. Further work will be needed to determine whether these events are causes of ALS but incompletely penetrant. We also identified many events that were previously unknown in cancer or healthy individuals. Most of these did not reach statistical significance, but that may reflect the relatively small number of control samples currently available.

The exact mechanisms leading to fusion events are not completely understood. Alterations in both DDR and RNA metabolism have been implicated as potential mechanisms leading to fusion events.^11^ Specifically, defects in DDR mechanisms have been described in motor neurons derived from people living with ALS and were associated with faster disease progression.^37^ Therefore, it is possible that alterations in DDR may be one possible mechanism leading to intra-chromosomal fusion events in ALS. In our study, many fusions unique to ALS involved non-coding genes and non-coding RNA (ncRNAs). Therefore, alterations in RNA splicing could also account for some of the intra-chromosomal fusions reported in this study, perhaps through the contribution of another rare phenomenon, trans-splicing.

The identification and characterization of fusion events in cancer has notably improved diagnosis, prognosis and treatment.^17, 21^ For example, the *CLDN18*--*ARHGAP* fusion is an important diagnostic and prognostic risk factor for gastric cancer,^38^ while the *DNAJB1*--*PRKACA* chimeric transcript contributes to the pathogenesis of the fibrolamellar carcinoma (FC).^19, 39^ Gene fusion events have been also described in brain cancers with several targetable fusion events in malignant gliomas^22, 28^ and neuroblastomas.^40^ Additionally, the identification of gene fusions has recently been applied to constitutional diseases, specifically in a variety of rare, undiagnosed phenotypes, which was found to result in improved diagnoses as well.^41–42^ As ALS is a multifactorial and heterogenous neurodegenerative disease arising from a combination of genetic and environmental factors, the enrichment of gene fusions we have identified here may suggest a role for structural genomic anomalies in ALS risk, onset or progression.

## Data Availability

All data produced in the present study are available upon reasonable request to the corresponding author.

## Author Contributions

Study design: YR, AAD, TP, EF, SMKF, and GSV. Study advisors: SEK, JDB, MEC, and KV. Data analysis: YR and AAD. Drafting of the manuscript: YR, AAD, TP, EF, SMKF, and GSV with input from all authors. Supervised the study: EF, SMKF, and GSV.

## Acknowledgements

The authors would like to thank the Target ALS Human Postmortem Tissue Core, New York Genome Center for Genomics of Neurodegenerative Disease, Amyotrophic Lateral Sclerosis Association and TOW Foundation. All NYGC ALS Consortium activities are supported by the ALS Association (ALSA, 19-Si-459) and the Tow Foundation. A.A.D. was supported by the Banting Postdoctoral Fellowships program. T.P. was supported by the Robert F. Schoeni Award for Research from Active Against ALS. The authors would like to thank Dr. Jack Humphrey from the Icahn School of Medicine at Mount Sinai for sharing his technical expertise during data analysis.

---

**Supplementary Table 1.** Demographics of tissue samples from ALS and controls. Sex was unknown for two control samples. Age at symptom onset was unknown for 53 ALS samples.

|  | **ALS** | **Control** |
| --- | --- | --- |
| **Total Subjects** | 367 | 90 |
| Target ALS Subjects | 177 | 10 |
| ALS Consortium Subjects | 177 | 80 |
| **Male:Female** | 200:167 | 52:36 |
| **Ethnicity (self-reported)** |  |  |
| Hispanic or Latino | 1 | 1 |
| Not Hispanic or Latino | 131 | 8 |
| Ashkenazi Jewish | 1 | 0 |
| Unknown | 227 | 80 |
| White Scottish | 7 | 1 |
| **Tissues Samples per Subject (mean; SD)** | 4.20 (2.26) | 2.77 (1.61) |
| **Tissue Samples** | 1542 | 249 |
| Motor Cortex | 373 | 48 |
| Spinal Cord – Cervical | 265 | 45 |
| Spinal Cord – Thoracic | 56 | 5 |
| Spinal Cord – Lumbar | 252 | 41 |
| Frontal Cortex | 248 | 48 |
| Sensory Cortex | 2 | 0 |
| Temporal Cortex | 34 | 25 |
| Occipital Cortex | 64 | 6 |
| Hippocampus | 61 | 13 |
| Cerebellum | 187 | 29 |
| **Capture Library Preparation Method** |  |  |
| Automated | 1047 | 203 |
| Manual | 495 | 46 |
| **Age at Symptom Onset (mean; SD)** | 60.52 (11.32) | N/A |
| **Bulbar Onset: Limb Onset: Other/Unknown** | 112:218:37 | N/A |

**Supplementary Table 2.** Top intra-chromosomal gene fusion pairs in each tissue displaying significant enrichment in ALS samples versus controls. The individual burden tests of gene fusion events were only performed on fusions within tissues demonstrating significant differences in the number of gene fusions of each intra-chromosomal subtype carried by ALS and control samples from each tissue type. Bonferroni and FDR corrections were based on the total number of fusions observed within the respective tissues. Abbreviations: ALS, amyotrophic lateral sclerosis; CI, confidence interval; FDR, false discovery rate; n, total number of samples; OR, odds ratio.

| Intra-chromosomal Fusion Type | Tissue Type | Fusion Name | ALS (n) | Control (n) | OR | Lower 95% CI | Upper 95% CI | p-value | Bonferroni p-value | FDR p-value |
| --- | --- | --- | --- | --- | --- | --- | --- | --- | --- | --- |
| Local Rearrangement | Motor Cortex | CAVIN4--TMEFF1 | 201 (373) | 12 (37) | 2.43 | 1.14 | 5.48 | 1.53E-02 | 1.00 | 1.00 |
|  |  | AL731769.2--SCART1 | 0 (373) | 1 (37) | 0.03 | 0.00 | 3.87 | 9.02E-02 | 1.00 | 1.00 |
|  |  | ERMP1--KIAA2026 | 28 (373) | 0 (37) | 6.19 | 0.72 | Inf | 9.49E-02 | 1.00 | 1.00 |
|  | Spinal Cord – Cervical | AC006427.2--TAPT1-AS1 | 118 (265) | 8 (45) | 3.70 | 1.62 | 9.55 | 8.59E-04 | 4.04E-02 | 4.04E-02 |
|  |  | AF241728.1--TSPAN7 | 0 (265) | 2 (45) | 0.03 | 0.00 | 0.89 | 2.07E-02 | 9.72E-01 | 4.86E-01 |
|  |  | CLIC2--AC234781.1 | 3 (265) | 3 (45) | 0.16 | 0.02 | 1.25 | 4.18E-02 | 1.00 | 6.55E-01 |
|  | Spinal Cord – Lumbar | AC006427.2--TAPT1-AS1 | 99 (252) | 4 (41) | 5.96 | 2.05 | 23.72 | 1.50E-04 | 7.48E-03 | 7.48E-03 |
|  |  | AC009975.1--STON1-GTF2A1L | 85 (252) | 5 (41) | 3.65 | 1.36 | 12.36 | 5.61E-03 | 2.80E-01 | 1.40E-01 |
|  |  | AC012414.4--AC012414.2 | 0 (252) | 1 (41) | 0.05 | 0.00 | 6.35 | 1.40E-01 | 1.00 | 1.00 |
|  |  | RANBP2--LIMS1 | 0 (252) | 1 (41) | 0.05 | 0.00 | 6.35 | 1.40E-01 | 1.00 | 1.00 |
|  |  | SMG8--PRR11 | 0 (252) | 1 (41) | 0.05 | 0.00 | 6.35 | 1.40E-01 | 1.00 | 1.00 |
|  | Frontal Cortex | AC006427.2--TAPT1-AS1 | 89 (248) | 6 (48) | 3.90 | 1.57 | 11.68 | 1.18E-03 | 9.07E-02 | 5.45E-02 |
|  |  | AC096642.2--LYPLAL1 | 38 (248) | 0 (48) | 17.74 | 2.14 | Inf | 1.45E-03 | 1.12E-01 | 5.45E-02 |
|  |  | CAVIN4--TMEFF1 | 111 (248) | 10 (48) | 3.07 | 1.42 | 7.22 | 2.12E-03 | 1.63E-01 | 5.45E-02 |
|  | Cerebellum | L3HYPDH--CCDC175 | 107 (187) | 7 (29) | 4.18 | 1.62 | 12.16 | 1.14E-03 | 9.84E-02 | 6.85E-02 |
|  |  | KPNA2P3--BPTFP1 | 89 (187) | 5 (29) | 4.33 | 1.53 | 15.17 | 2.22E-03 | 1.91E-01 | 6.85E-02 |
|  |  | AC009975.1--STON1-GTF2A1L | 52 (187) | 1 (29) | 10.72 | 1.68 | 448.50 | 2.39E-03 | 2.05E-01 | 6.85E-02 |
| Not Close Proximity | Cerebellum | ABCD2--KIF21A | 66 (182) | 2 (28) | 7.35 | 1.75 | 65.94 | 1.85E-03 | 6.19E-02 | 6.19E-02 |
|  |  | PMS2P11--CCDC146 | 0 (182) | 2 (28) | 0.03 | 0 | 0.80 | 1.72E-02 | 9.99E-01 | 5.00E-01 |
|  |  | AC023421.2--AC021517.2 | 0 (182) | 1 (28) | 0.05 | 0 | 6.00 | 1.33E-01 | 1.00 | 1.00 |
|  |  | DOCK4--IMMP2L | 0 (182) | 1 (28) | 0.05 | 0 | 6.00 | 1.33E-01 | 1.00 | 1.00 |
|  |  | PCSK6--TARS3 | 0 (182) | 1 (28) | 0.05 | 0 | 6.00 | 1.33E-01 | 1.00 | 1.00 |
| Neighbor | Spinal Cord – Cervical | PAMR1--SLC1A2 | 56 (265) | 0 (45) | 24.54 | 3.00 | Inf | 8.94E-05 | 8.50E-03 | 8.50E-03 |
|  |  | C8orf44--SGK3 | 42 (265) | 1 (45) | 8.26 | 1.33 | 342.05 | 9.86E-03 | 9.37E-01 | 4.51E-01 |
|  |  | MAILR--ATP6V1C1 | 60 (265) | 3 (45) | 4.08 | 1.24 | 21.32 | 1.48E-02 | 1.00 | 4.51E-01 |
|  | Spinal Cord – Lumbar | TVP23C--CDRT4 | 136 (252) | 11 (41) | 3.19 | 1.47 | 7.37 | 1.32E-03 | 1.17E-01 | 1.17E-01 |
|  |  | IRF1-AS1--RAD50 | 73 (252) | 5 (41) | 2.93 | 1.08 | 9.94 | 2.29E-02 | 1.00 | 7.33E-01 |
|  |  | AC012405.1--AC073941.1 | 80 (252) | 6 (41) | 2.71 | 1.07 | 8.19 | 2.66E-02 | 1.00 | 7.33E-01 |
|  | Frontal Cortex | AC090517.5--ZNF280D | 191 (248) | 26 (48) | 2.82 | 1.41 | 5.63 | 2.10E-03 | 2.16E-01 | 1.61E-01 |
|  |  | TVP23C--CDRT4 | 166 (248) | 21 (48) | 2.59 | 1.32 | 5.15 | 3.12E-03 | 3.21E-01 | 1.61E-01 |
|  |  | SH3GL3--ADAMTSL3 | 30 (248) | 0 (48) | 13.54 | 1.61 | Inf | 6.98E-03 | 7.19E-01 | 2.40E-01 |
|  | Temporal Cortex | AEBP2--AC024901.1 | 13 (34) | 0 (25) | 32.02 | 3.08 | Inf | 2.74E-04 | 1.12E-02 | 1.12E-02 |
|  |  | TVP23C--CDRT4 | 23 (34) | 6 (25) | 6.38 | 1.82 | 25.57 | 1.43E-03 | 5.87E-02 | 2.93E-02 |
|  |  | AC090517.5--ZNF280D | 23 (34) | 8 (25) | 4.32 | 1.30 | 15.64 | 8.98E-03 | 3.68E-01 | 1.23E-01 |
|  | Hippocampus | EFCAB2--KIF26B | 0 (61) | 3 (13) | 0.04 | 0 | 0.47 | 4.41E-03 | 2.12E-01 | 2.12E-01 |
|  |  | TVP23C--CDRT4 | 36 (61) | 4 (13) | 3.19 | 0.78 | 15.78 | 7.49E-02 | 1.00 | 1.00 |
|  |  | MAILR--ATP6V1C1 | 18 (61) | 1 (13) | 4.94 | 0.64 | 226.24 | 1.63E-01 | 1.00 | 1.00 |
|  | Cerebellum | EEF1AKNMT--DNM3 | 50 (187) | 2 (29) | 4.90 | 1.16 | 44.06 | 1.91E-02 | 1.00 | 1.00 |
|  |  | AC090517.5--ZNF280D | 92 (187) | 9 (29) | 2.14 | 0.88 | 5.64 | 7.49E-02 | 1.00 | 1.00 |
|  |  | OTUB1--AP000721.2 | 0 (187) | 1 (29) | 0.05 | 0.00 | 6.05 | 1.34E-01 | 1.00 | 1.00 |
| Overlapping Neighbor | Motor Cortex | AC127502.3--AC127502.1 | 52 (373) | 2 (37) | 2.83 | 0.69 | 25.00 | 2.02E-01 | 1.00 | 1.00 |
|  |  | KHDRBS2-OT1--KHDRBS2 | 371 (373) | 36 (37) | 5.12 | 0.09 | 100.56 | 2.48E-01 | 1.00 | 1.00 |
|  |  | Z68871.1--LINC00630 | 325 (373) | 30 (37) | 1.58 | 0.55 | 3.94 | 3.12E-01 | 1.00 | 1.00 |
|  | Frontal Cortex | AC067956.1--AC019211.1 | 71 (248) | 4 (48) | 4.40 | 1.52 | 17.48 | 1.97E-03 | 4.34E-02 | 4.34E-02 |
|  |  | AC127502.3--AC127502.1 | 39 (248) | 2 (48) | 4.28 | 1.04 | 37.89 | 3.81E-02 | 8.38E-01 | 4.19E-01 |
|  |  | LINC02263--LINC01378 | 2 (248) | 1 (48) | 0.38 | 0.02 | 23.02 | 4.13E-01 | 1.00 | 1.00 |

**Supplementary Figure 1.**
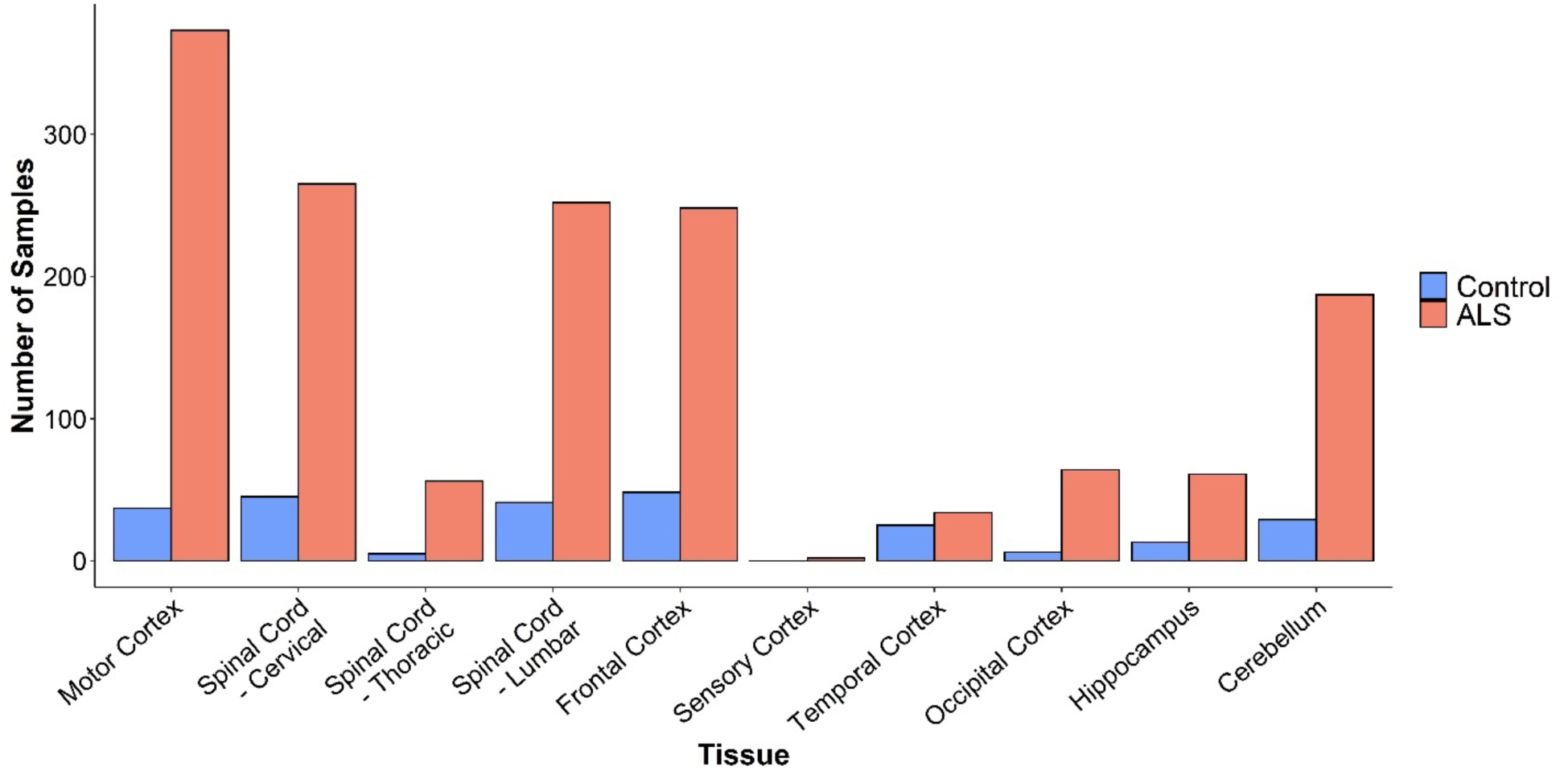
Distribution of tissues across ALS and control samples. Several brain regions as well as spinal cord regions were collected per individual ALS and control.

**Supplementary Figure 2.**
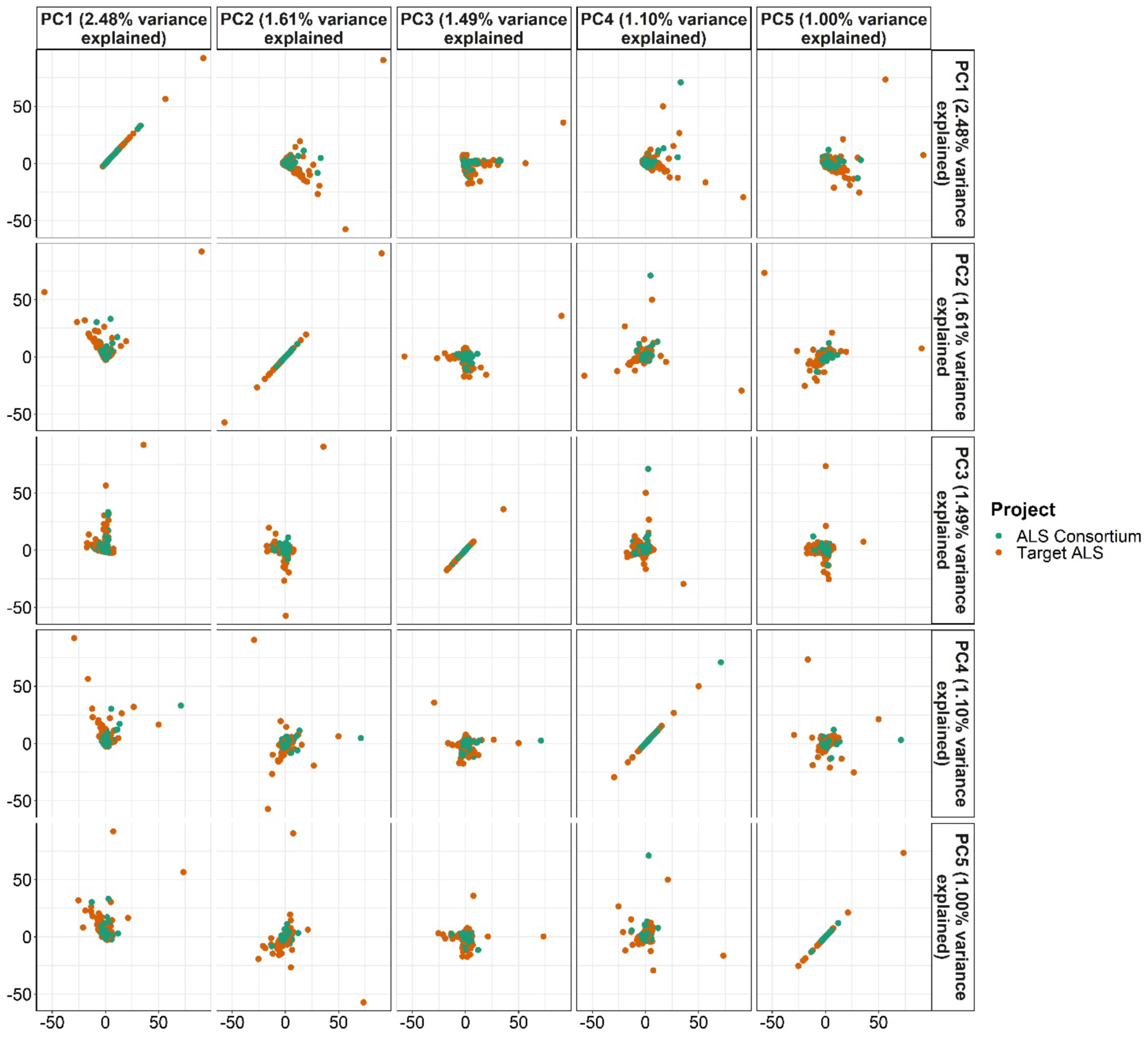
Assessment of any possible batch effects of Target ALS and ALS Consortium gene fusion data using PCA of fusion concentrations. PCA analysis was applied upon a matrix of fusion fragments per million total RNA-Seq fragments (FFPM) values. The first five principal components are displayed in this figure.

**Supplementary Figure 3.**
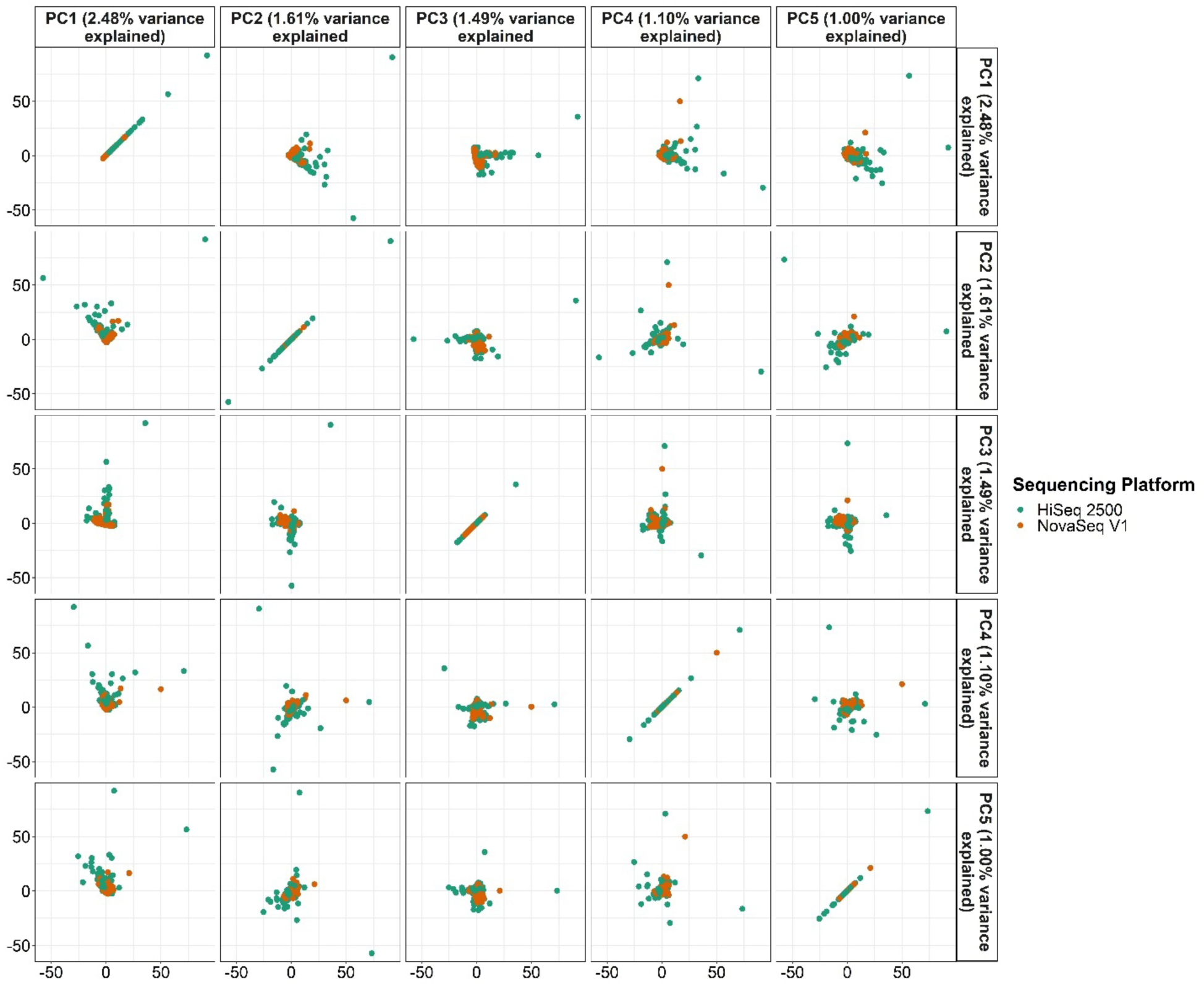
Assessment of any possible batch effects of the sequencing platform used for RNA-Seq of the samples using PCA of fusion concentrations. PCA analysis was applied upon a matrix of fusion fragments per million total RNA-Seq fragments (FFPM) values. The first five principal components are displayed in this figure.

**Supplementary Figure 4.**
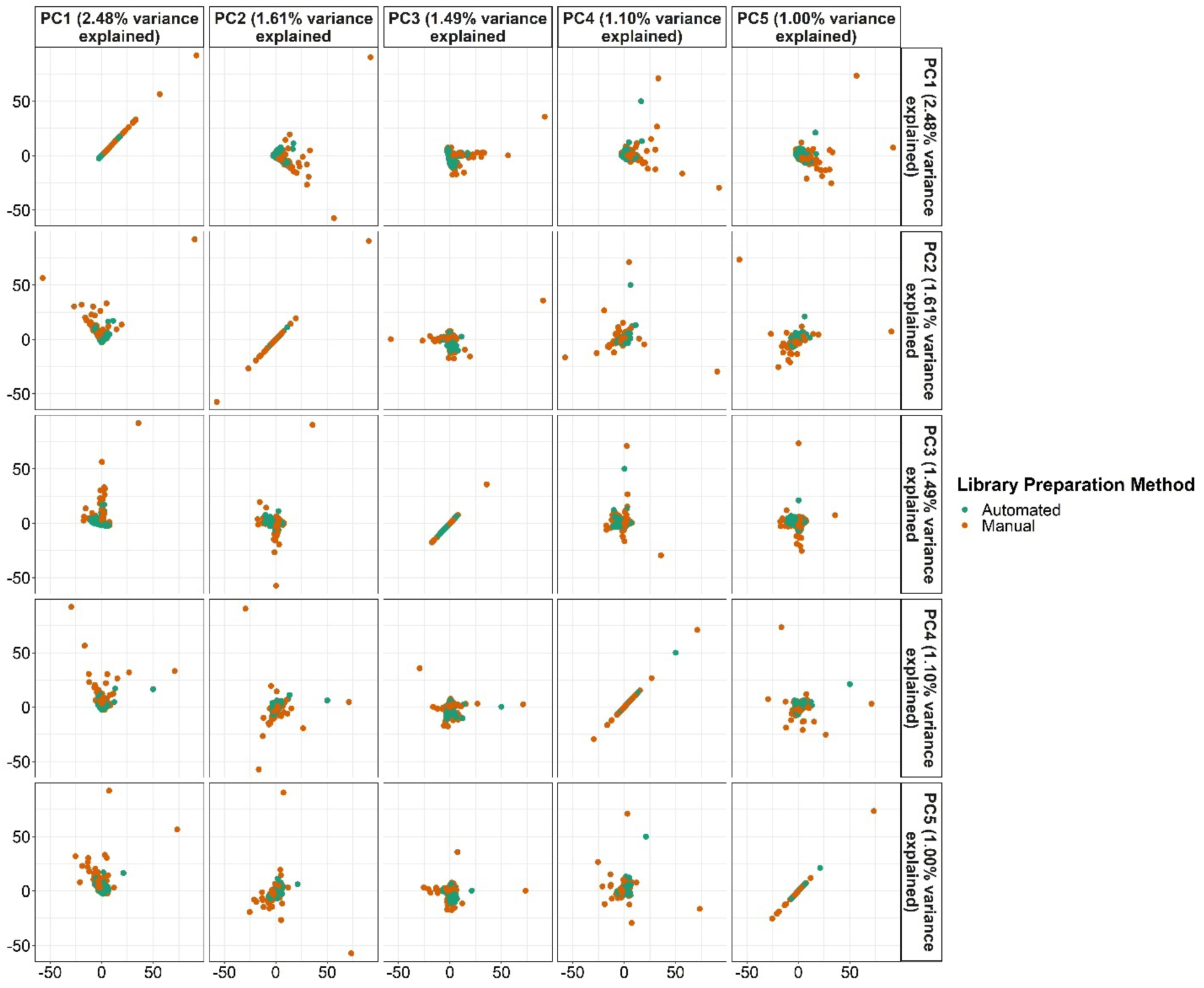
Assessment of any possible batch effects of sample capture library preparation method using PCA of fusion concentrations. PCA analysis was applied upon a matrix of fusion fragments per million total RNA-Seq fragments (FFPM) values. The first five principal components are displayed in this figure.

**Supplementary Figure 5.**
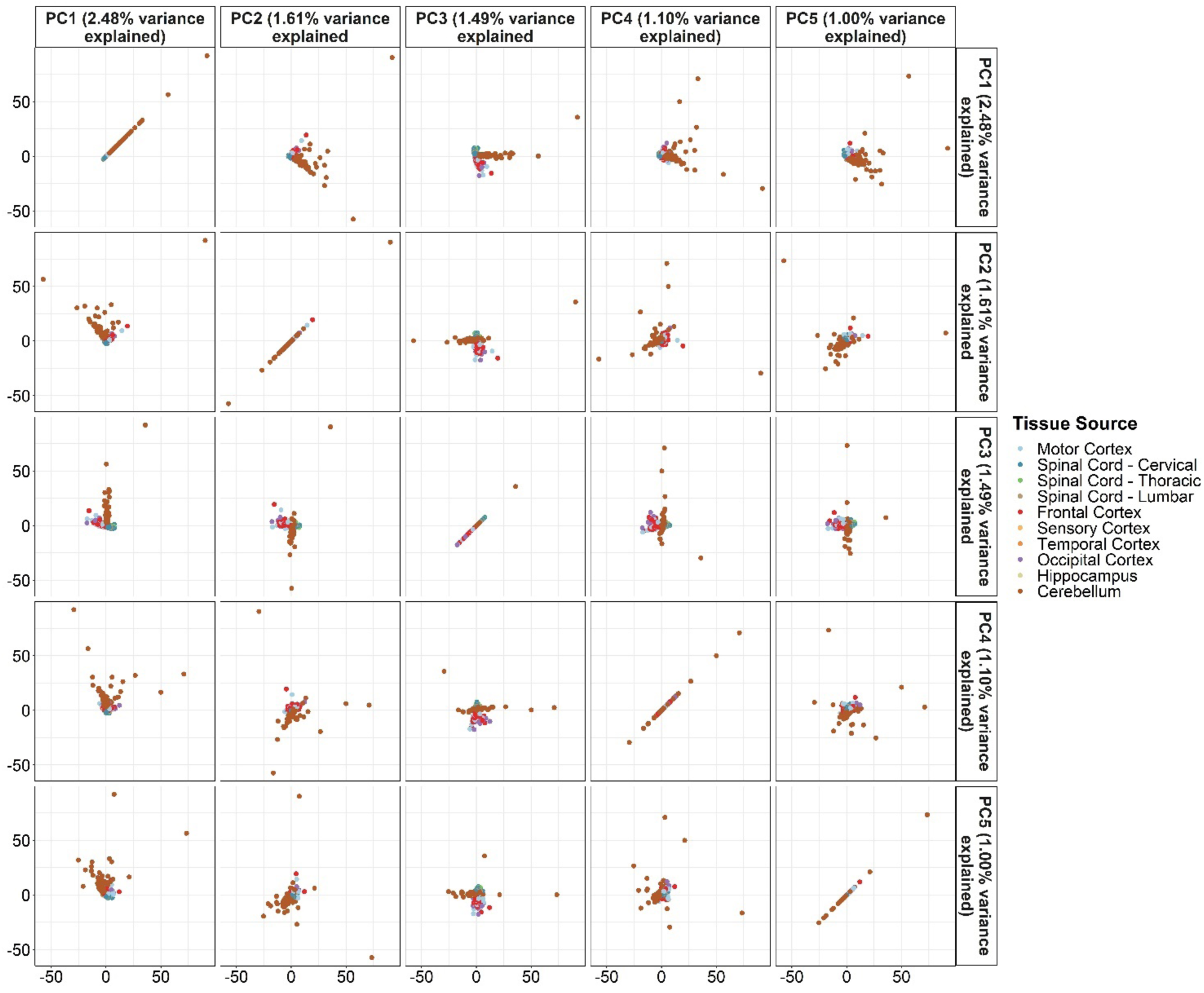
Assessment of any possible batch effects of sample tissue of origin using PCA of fusion concentrations. PCA analysis was applied upon a matrix of fusion fragments per million total RNA-Seq fragments (FFPM) values. The first five principal components are displayed in this figure.

**Supplementary Figure 6.**
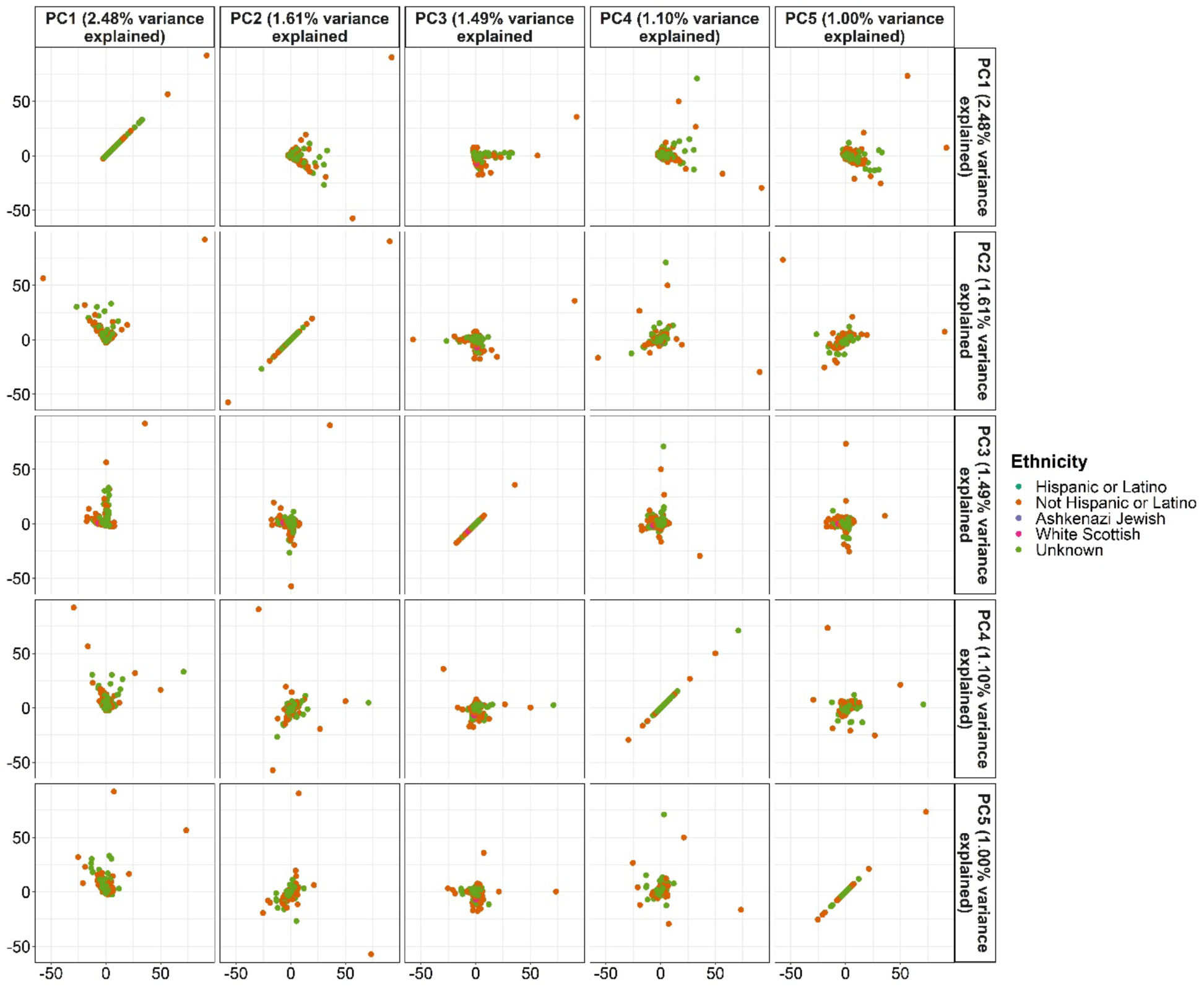
Assessment of any possible batch effects of ethnicity of the subject from which the sample was obtained using PCA of fusion concentrations. PCA analysis was applied upon a matrix of fusion fragments per million total RNA-Seq fragments (FFPM) values. The first five principal components are displayed in this figure.

**Supplementary Figure 7.**
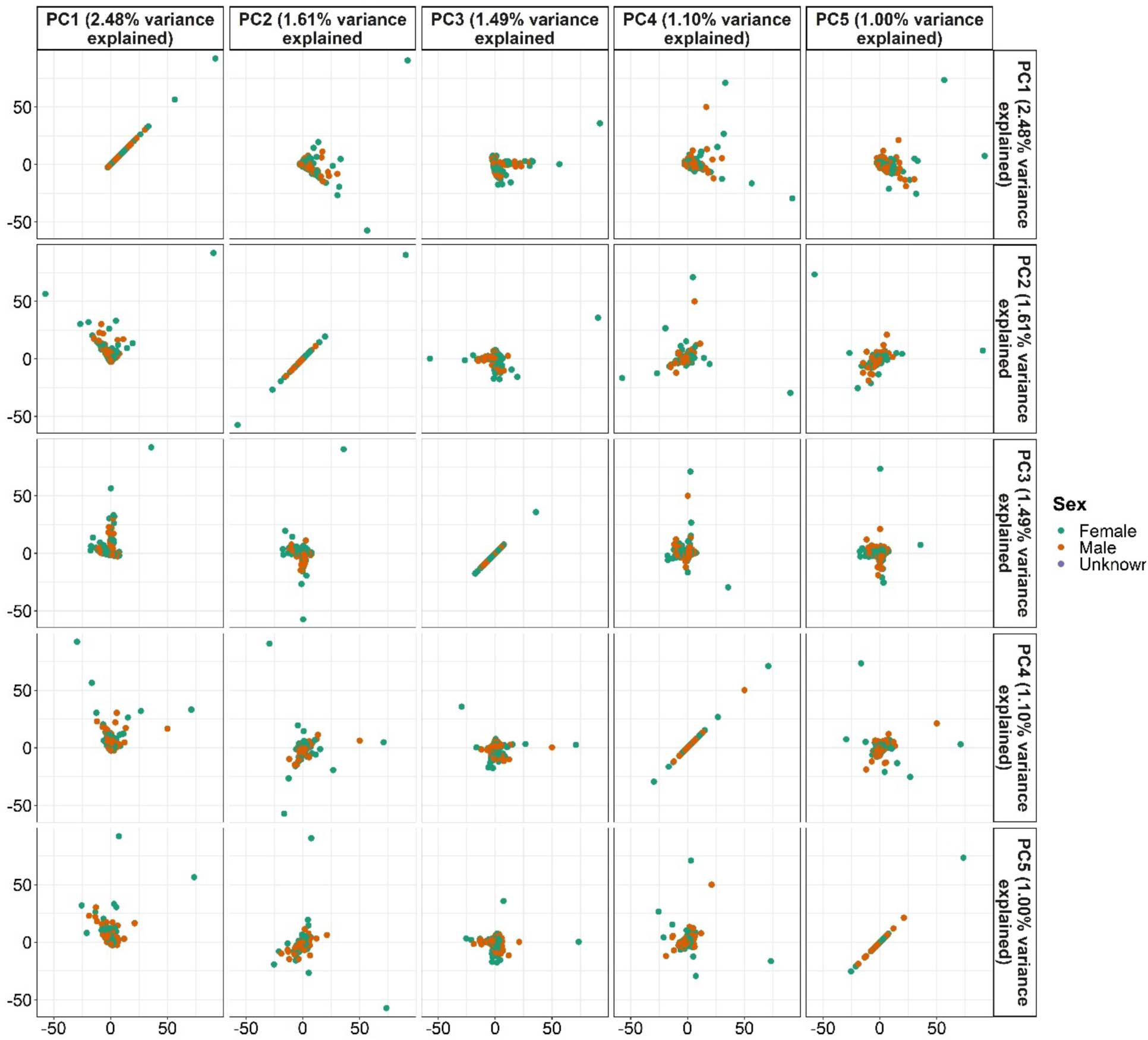
Assessment of any possible batch effects of sex of the subject from which the sample was obtained using PCA of fusion concentrations. PCA analysis was applied upon a matrix of fusion fragments per million total RNA-Seq fragments (FFPM) values. The first five principal components are displayed in this figure.

**Supplementary Figure 8.**
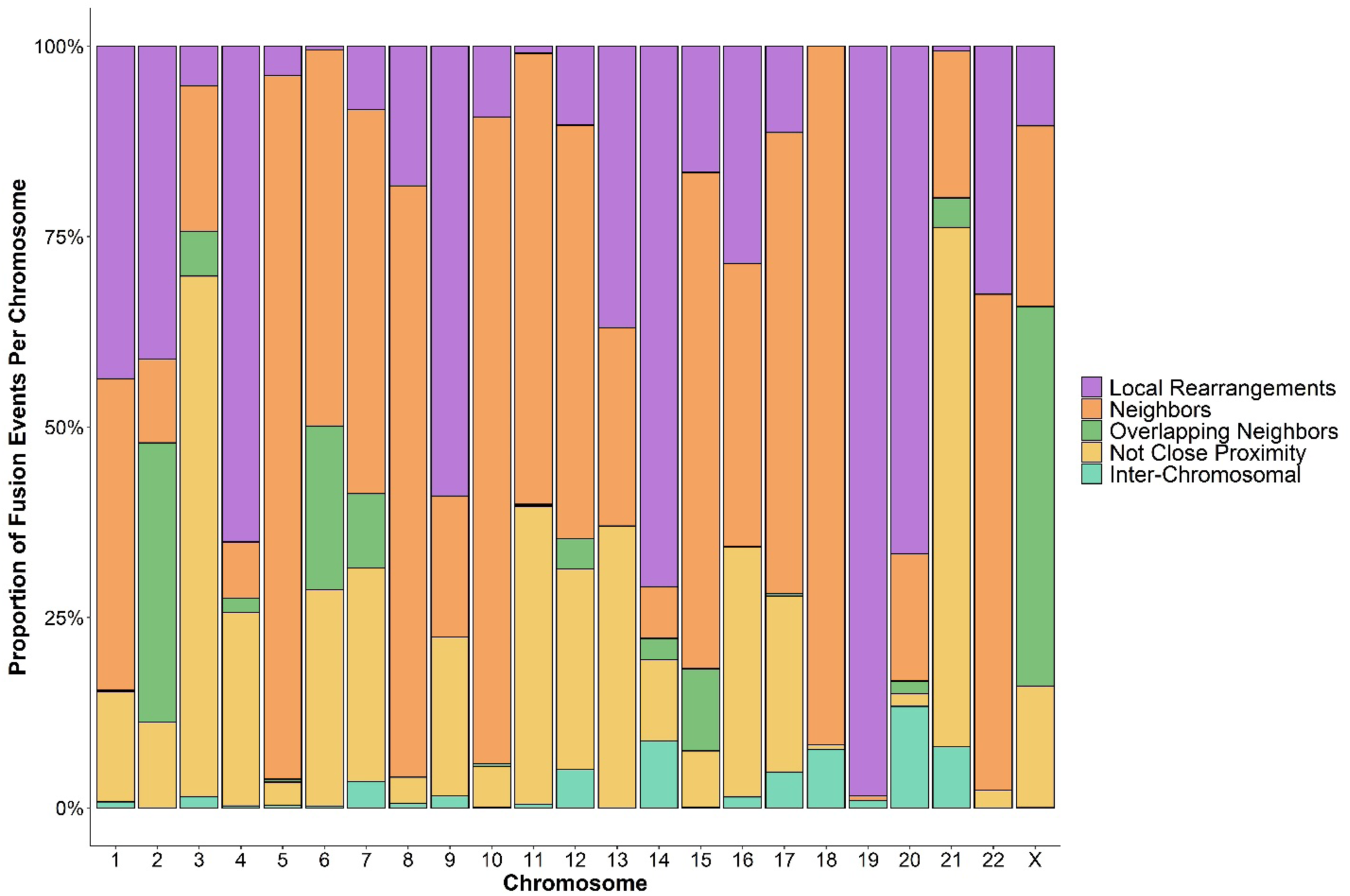
Proportion of gene fusions of each subtype per chromosome. The proportion of breakpoint-unique gene fusions in both ALS and control samples encompassed by each chromosome based on subtypes which included both intra-chromosomal fusions (local rearrangements, neighbor fusions, overlapping neighbor, and not close proximity) and inter-chromosomal fusions.

**Supplementary Figure 9.**
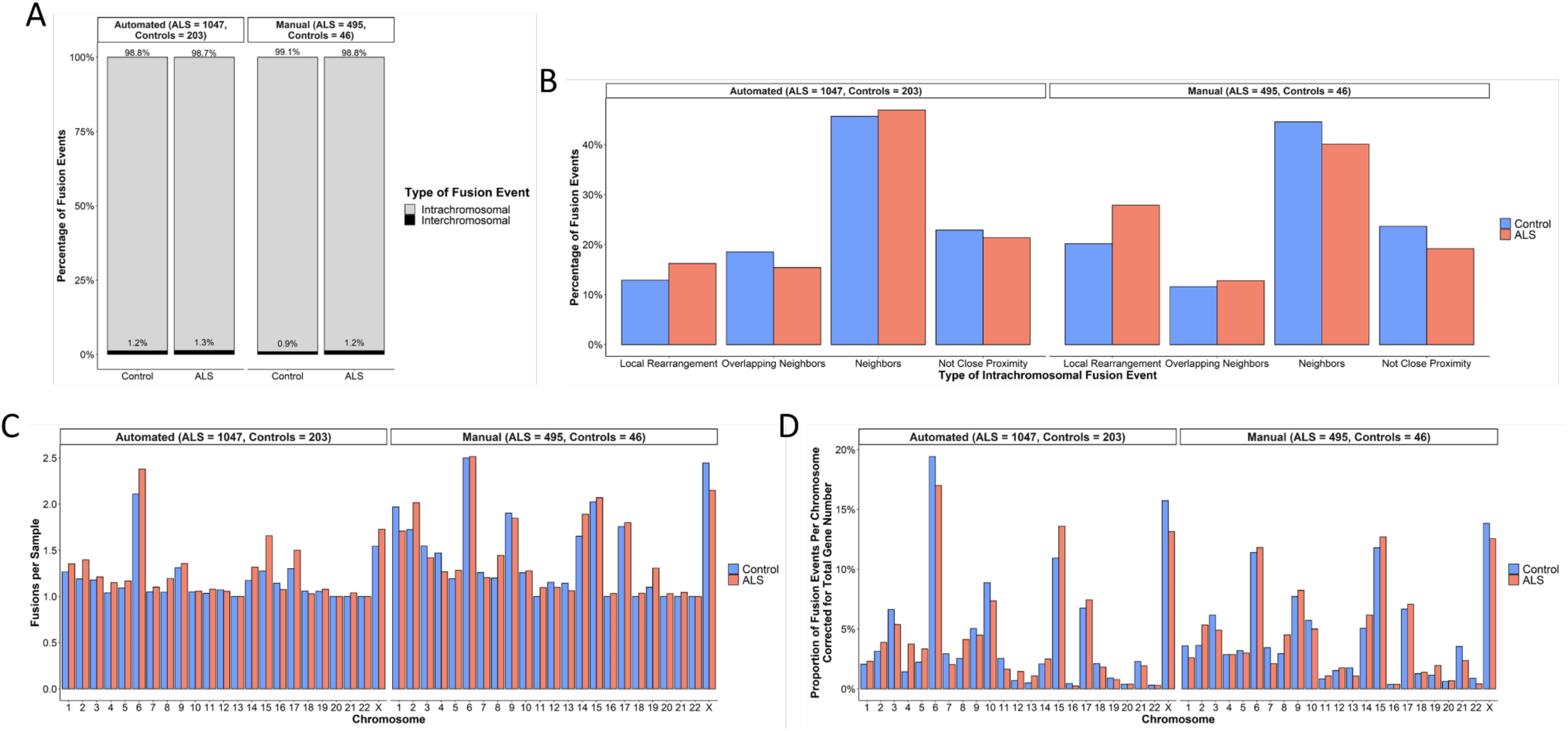
Characterization of the breakpoint-unique gene fusions and their subtypes in ALS and control samples subdivided by capture library preparation method. The breakpoint-unique gene fusions in ALS and control samples subdivided by capture library preparation method were compared to determine the distribution of (A) intra-chromosomal and inter-chromosomal gene fusions, (B) intra-chromosomal gene fusion subtypes, (C) fusion events per sample based on the chromosome(s) involved, and (D) the proportion of fusion events per chromosome(s) involved in the gene fusions corrected for total number of genes located on the chromosome (Ensembl, release 106).

**Supplementary Figure 10.**
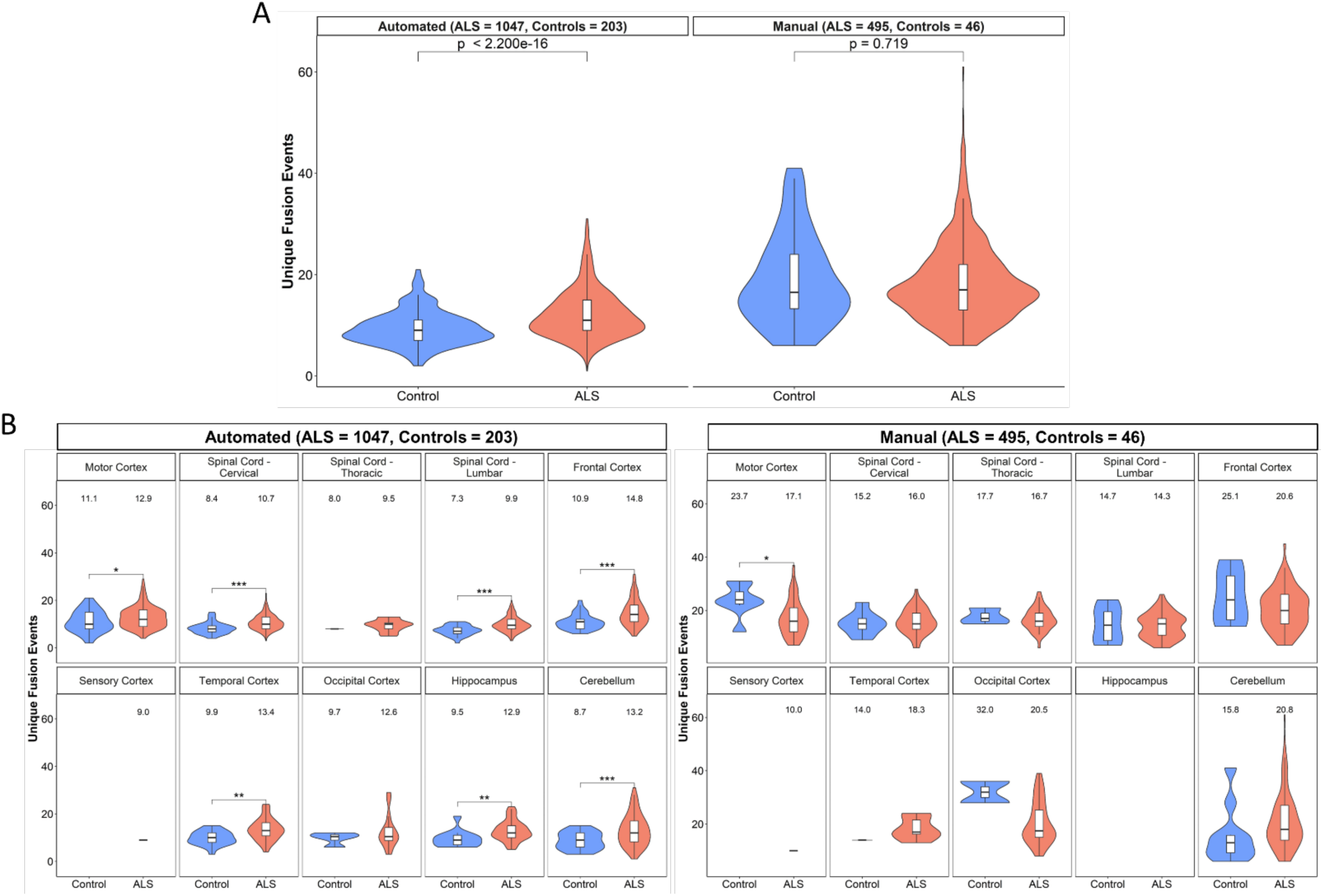
Breakpoint-unique gene fusions carried in ALS and control samples subdivided by capture library preparation method. The distribution of breakpoint-unique gene fusions carried per sample was compared between ALS and control samples using the Welch’s t-test, both independent of sample tissue source and within each individual tissue source. (A) ALS samples prepared with the automated library preparation method carried significantly more breakpoint-unique gene fusions (mean = 12.18, sd = 4.72) than control samples (mean = 9.41, sd = 3.41), but not ALS samples prepared with the manual library preparation method (ALS samples: mean = 18.43, sd = 7.72; control samples: mean = 18.91, sd = 8.82). (B) Significantly more breakpoint-unique gene fusions were carried by ALS samples compared to controls prepared with the automated library preparation method in the motor cortex (p = 0.0457, cervical spinal cord (p = 3.972e-06), lumbar spinal cord (p = 5.523e-08), frontal cortex (p = 4.221e-08), temporal cortex (p = 1.839e-4), hippocampus (p = 0.0056), and cerebellum (p = 2.625e-4). Significantly fewer breakpoint-unique gene fusions were carried by ALS samples compared to controls prepared with the manual library preparation method in the motor cortex (p = 0.0314). No statistical comparisons were performed in the thoracic spinal cord, sensory cortex, or occipital cortex as there were too few (n < 10) tissue samples from controls. * < 0.05; ** < 0.01; *** p < 0.001.

**Supplementary Figure 11.**
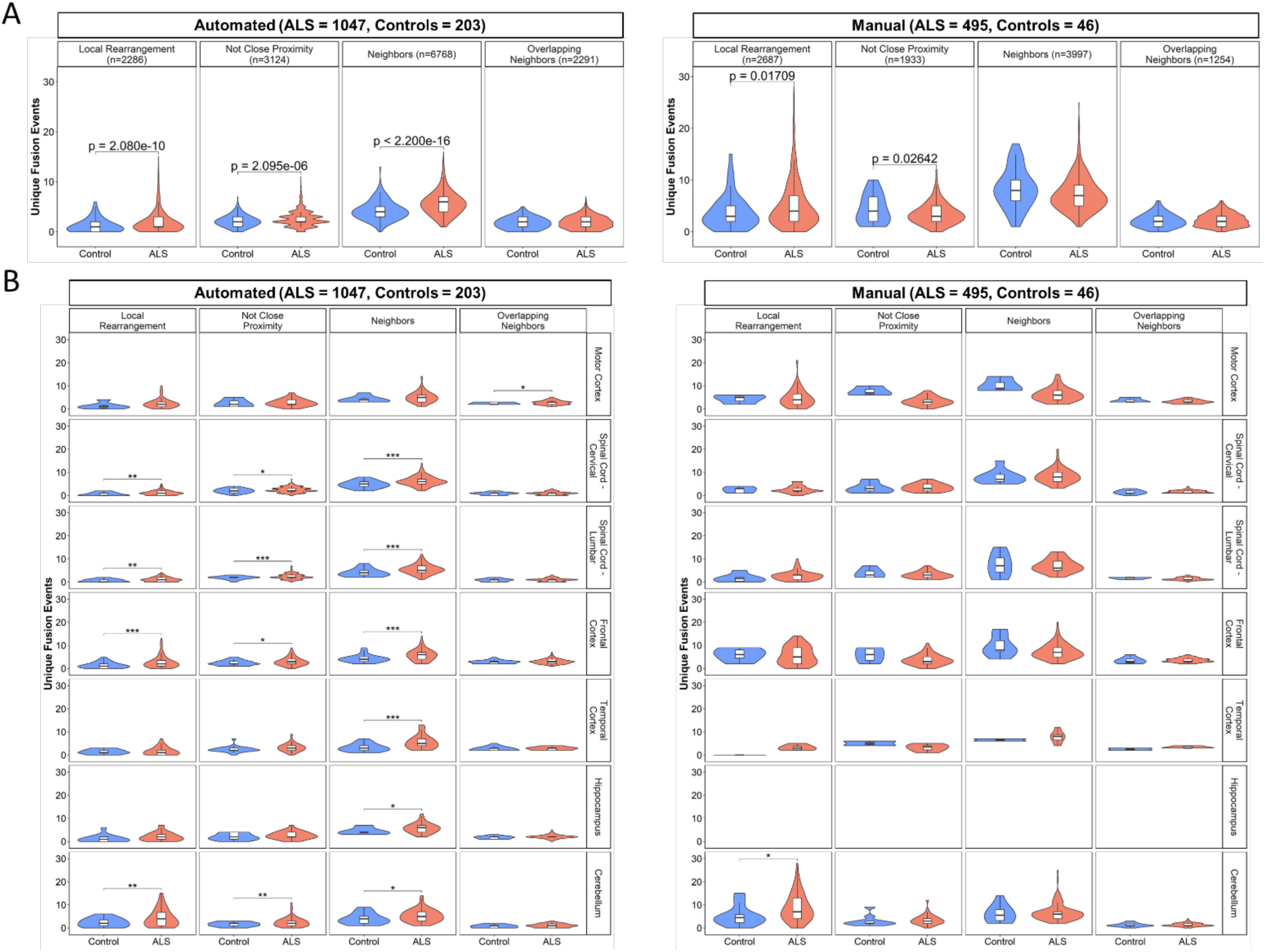
Enrichment of breakpoint-unique gene fusions carried by ALS and control samples subdivided by capture library preparation method. Welch’s t-test was used to compare the number of breakpoint-unique gene fusions of each subtype carried by each ALS and control sample both independent of sample tissue source and within each individual tissue source. (A) There was a significant enrichment of the intra chromosomal subtypes local rearrangements, not close proximity fusions, and neighbor fusions in ALS samples compared to control samples prepared using automated capture library preparation methods. There was a significant enrichment of the intra chromosomal subtypes local rearrangements and not close proximity fusions in ALS samples compared to control samples prepared using manual capture library preparation methods. (B) Following subgrouping of gene fusion based on the tissue source of the sample in which they were identified, a significant over-representation of local rearrangement events was identified in ALS samples from the cervical spinal cord (p = 0.0018), lumbar spinal cord (p = 0.0017), frontal cortex (p = 3.672e-06), and cerebellum (p = 0.0024) in ALS samples compared to control samples prepared with automated methods. Not close proximity gene fusion events were significantly enriched in ALS samples prepared with automated methods from the cervical spinal cord (p = 0.0219), lumbar spinal cord (p = 8.637e-04), frontal cortex (p = 0.0170), and cerebellum (p = 0.0075). Neighbor gene fusion events were significantly enriched in the ALS samples prepared with automated methods from the cervical spinal cord (p = 4.614e-06), lumbar spinal cord (p = 4.543e-05), frontal cortex (p = 1.130e-06), temporal cortex (p = 9.118e-04), hippocampus (p = 0.0155), and cerebellum (p = 0.0477). Overlapping neighbor gene fusion events were significantly enriched in ALS samples prepared with automated methods from the motor cortex (p = 0.0146). Local rearrangement gene fusion events were significantly enriched in ALS samples prepared with manual methods from the cerebellum (p = 0.0381). No enrichment analyses were performed in the thoracic spinal cord, sensory cortex, or occipital cortex as there were too few (n < 10) tissue samples from controls. * < 0.05; ** < 0.01; *** p < 0.001.

**Supplementary Figure 12.**
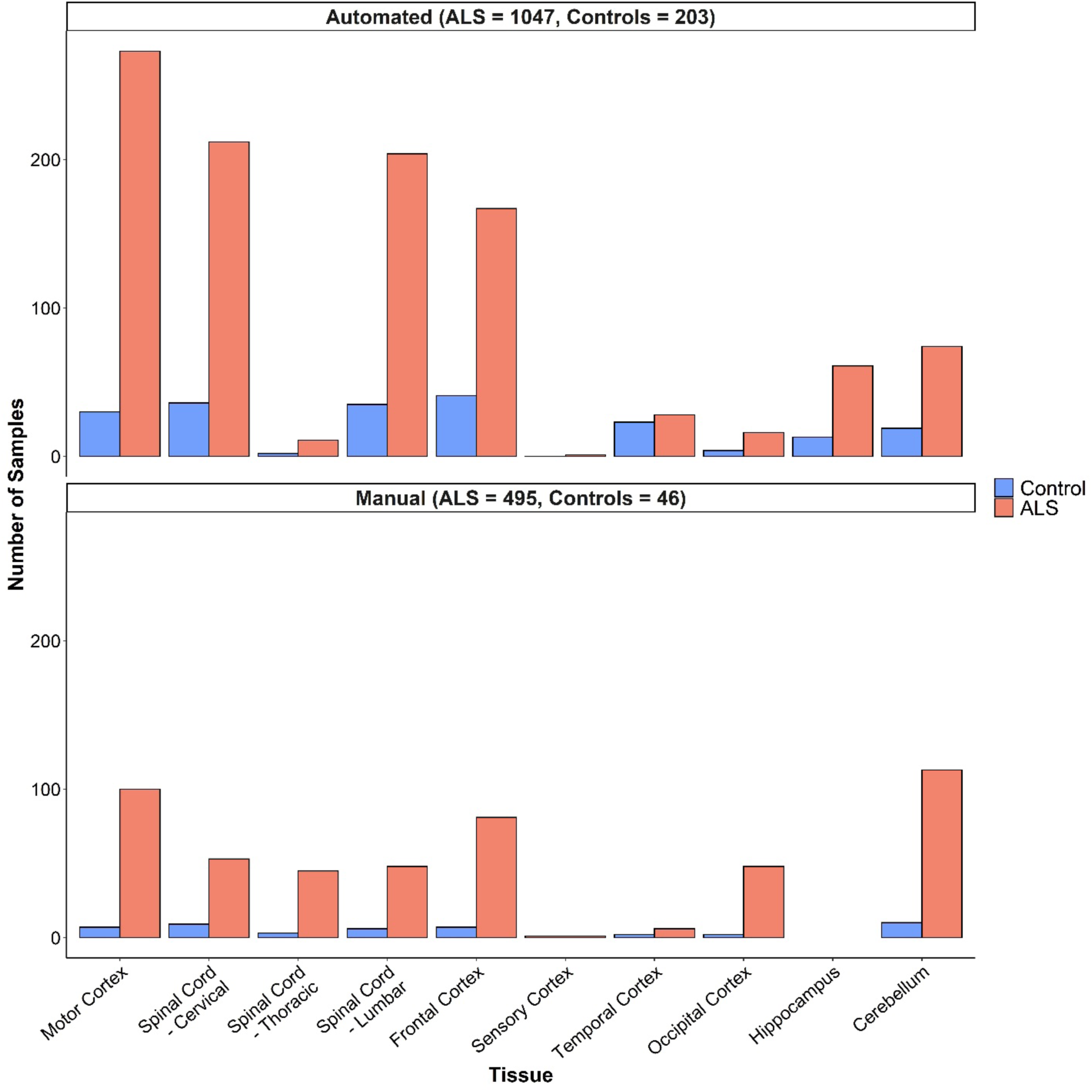
Distribution of tissues across ALS and control samples subdivided by capture library preparation method. Several brain regions as well as spinal cord regions were collected per individual ALS and control.

**Supplementary Figure 13.**
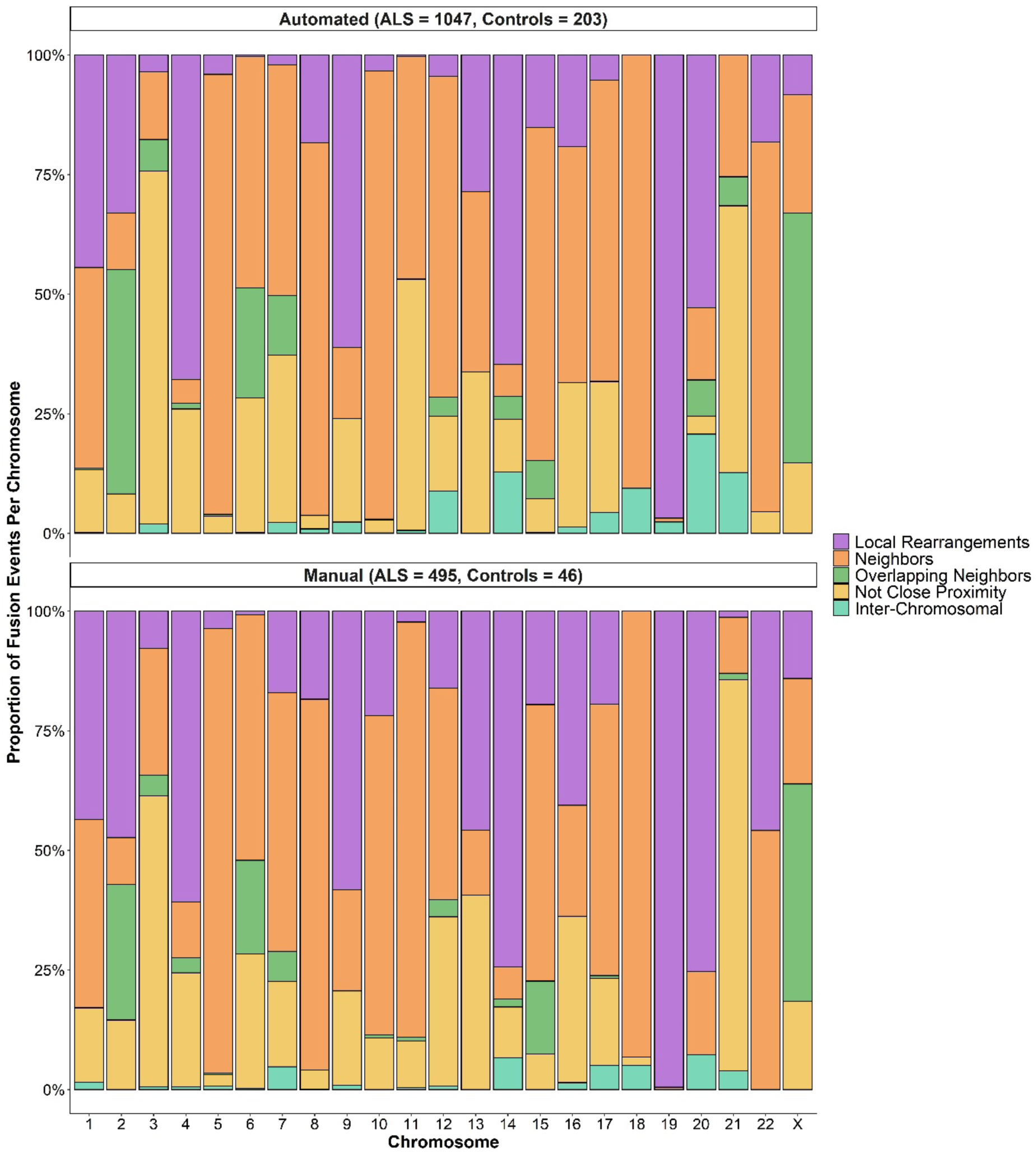
Proportion of gene fusions of each subtype per chromosome. The proportion of breakpoint-unique gene fusions in both ALS and control samples subdivided by capture library preparation method encompassed by each chromosome based on subtypes which included both intra-chromosomal fusions (local rearrangements, neighbor fusions, overlapping neighbor, and not close proximity) and inter-chromosomal fusions.

## Supplemental Materials

NYGC ALS Consortium

Hemali Phatnani^5^, Justin Kwan^6^, Dhruv Sareen^7,8^, James R. Broach^9^, Zachary Simmons^10^, Ximena Arcila-Londono^11^, Edward B. Lee^12^, Vivianna M. Van Deerlin^12^, Neil A. Shneider^13^, Ernest Fraenkel^1^, Lyle W. Ostrow^14^, Frank Baas^15,16^, Noah Zaitlen^17^, James D. Berry^18,19^, Andrea Malaspina^19,20,21^, Pietro Fratta^22^, Gregory A. Cox^23^, Leslie M. Thompson^24,25^, Steve Finkbeiner^26^, Efthimios Dardiotis^27^, Timothy M. Miller^28^, Siddharthan Chandran^29^, Suvankar Pal^29^, Eran Hornstein^30^, Daniel J. MacGowan^31^, Terry Heiman-Patterson^32^, Molly G. Hammell^33^, Nikolaos. A. Patsopoulos^34,35^, Oleg Butovsky^36^, Joshua Dubnau^37^, Avindra Nath^38^, Robert Bowser^39,40^, Matthew Harms^41^, Eleonora Aronica^42^, Mary Poss^43^, Jennifer Phillips-Cremins^44^, John Crary^45^, Nazem Atassi^46^, Dale J. Lange^47,48^, Darius J. Adams^49,50^, Leonidas Stefanis^51,52^, Marc Gotkine^53^, Robert H. Baloh^54,55^, Suma Babu^19^, Towfique Raj^56^, Sabrina Paganoni^57^, Ophir Shalem^58,59^, Colin Smith^60,61^, Bin Zhang^62^, Brent Harris^63^, Iris Broce^64^, Vivian Drory^65^, John Ravits^66^, Corey McMillan^67^, Vilas Menon^68^, Lani Wu^69^, Steven Altschuler^69^, Yossef Lerner^70^, Rita Sattler^71^, Kendall Van Keuren-Jensen^72^, Orit Rozenblatt-Rosen^73^, Kerstin Lindblad-Toh^73^, Katharine Nicholson^74^, Peter Gregersen^75^, Jeong-Ho Lee^76^, Sulev Kokos^77^, Stephen Muljo^78^, & Bryan J. Traynor^79^.

^5^Center for Genomics of Neurodegenerative Disease (CGND), New York Genome Center, New York, NY, USA. ^6^Department of Neurology, Lewis Katz School of Medicine, Temple University, Philadelphia, PA, USA. ^7^Cedars-Sinai Department of Biomedical Sciences, Board of Governors Regenerative Medicine Institute and Brain Program, Cedars-Sinai Medical Center, University of California, Los Angeles, CA, USA. ^8^Department of Medicine, University of California, Los Angeles, CA, USA. ^9^Department of Biochemistry and Molecular Biology, Penn State Institute for Personalized Medicine, The Pennsylvania State University, Hershey, PA, USA. ^10^Department of Neurology, The Pennsylvania State University, Hershey, PA, USA. ^11^Department of Neurology, Henry Ford Health System, Detroit, MI, USA. ^12^Department of Pathology and Laboratory Medicine, Perelman School of Medicine, University of Pennsylvania, Philadelphia, PA, USA. ^13^Department of Neurology, Center for Motor Neuron Biology and Disease, Institute for Genomic Medicine, Columbia University, New York, NY, USA. ^1^Department of Biological Engineering, Massachusetts Institute of Technology, Cambridge, MA, USA. ^14^Department of Neurology, Johns Hopkins School of Medicine, Baltimore, MD, USA. ^15^Department of Neurogenetics, Academic Medical Centre, Amsterdam, The Netherlands. ^16^Leiden University Medical Center, Leiden, The Netherlands. ^17^Department of Medicine, Lung Biology Center, University of California, San Francisco, CA, USA. ^18^ALS Multidisciplinary Clinic, Neuromuscular Division, Department of Neurology, Harvard Medical School, Boston, MA, USA. ^19^Neurological Clinical Research Institute, Massachusetts General Hospital, Boston, MA, USA. ^19^Centre for Neuroscience and Trauma, Blizard Institute, Barts, Queen Mary University of London, London, UK. ^20^The London School of Medicine and Dentistry, Queen Mary University of London, London, UK. ^21^Department of Neurology, Basildon University Hospital, Basildon, UK. ^22^Institute of Neurology, National Hospital for Neurology and Neurosurgery, University College London, London, UK. ^23^The Jackson Laboratory, Bar Harbor, ME, USA. ^24^Department of Psychiatry and Human Behavior, Department of Biological Chemistry, School of Medicine, University of California, Irvine, CA, USA. ^25^Department of Neurobiology and Behavior, School of Biological Sciences, University of California, Irvine, CA, USA. ^26^Taube/Koret Center for Neurodegenerative Disease Research, Roddenberry Center for Stem Cell Biology and Medicine, Gladstone Institute, San Francisco, CA, USA. ^27^Department of Neurology and Sensory Organs, University of Thessaly, Thessaly, Greece. ^28^Department of Neurology, Washington University in St Louis, St Louis, MO, USA. ^29^Centre for Clinical Brain Sciences, Anne Rowling Regenerative Neurology Clinic, Euan MacDonald Centre for Motor Neurone Disease Research, University of Edinburgh, Edinburgh, UK. ^30^Department of Molecular Genetics, Weizmann Institute of Science, Rehovot, Israel. ^31^Department of Neurology, Icahn School of Medicine at Mount Sinai, New York, NY, USA. ^32^Center for Neurodegenerative Disorders, Department of Neurology, the Lewis Katz School of Medicine, Temple University, Philadelphia, PA, USA. ^33^Cold Spring Harbor Laboratory, Cold Spring Harbor, NY, USA. ^34^Computer Science and Systems Biology Program, Ann Romney Center for Neurological Diseases, Department of Neurology and Division of Genetics in Department of Medicine, Brigham and Women’s Hospital, Harvard Medical School, Boston, MA, USA. ^35^Program in Medical and Population Genetics, Broad Institute, Cambridge, MA, USA. ^36^Ann Romney Center for Neurologic Diseases, Brigham and Women’s Hospital, Harvard Medical School, Boston, MA, USA. ^37^Department of Anesthesiology, Stony Brook University, Stony Brook, NY, USA. ^38^Section of Infections of the Nervous System, National Institute of Neurological Disorders and Stroke, NIH, Bethesda, MD, USA. ^39^Department of Neurology, Barrow Neurological Institute, St Joseph’s Hospital, Phoenix, AZ, USA. ^40^Medical Center, Department of Neurobiology, Barrow Neurological Institute, St Joseph’s Hospital and Medical Center, Phoenix, AZ, USA. ^41^Department of Neurology, Division of Neuromuscular Medicine, Columbia University, New York, NY, USA. ^42^Department of Neuropathology, Academic Medical Center, University of Amsterdam, Amsterdam, The Netherlands. ^43^Department of Biology and Veterinary and Biomedical Sciences, The Pennsylvania State University, University Park, PA, USA. ^44^New York Stem Cell Foundation, Department of Bioengineering, School of Engineering and Applied Sciences, University of Pennsylvania, Philadelphia, PA, USA. ^45^Department of Pathology, Fishberg Department of Neuroscience, Friedman Brain Institute, Ronald M. Loeb Center for Alzheimer’s Disease, Icahn School of Medicine at Mount Sinai, New York, NY, USA. ^46^Department of Neurology, Harvard Medical School, Neurological Clinical Research Institute, Massachusetts General Hospital, Boston, MA, USA. ^47^Department of Neurology, Hospital for Special Surgery, New York, NY, USA. ^48^Weill Cornell Medical Center, New York, NY, USA. ^49^Medical Genetics, Atlantic Health System, Morristown Medical Center, Morristown, NJ, USA. ^50^Overlook Medical Center, Summit, NJ, USA. ^51^Center of Clinical Research, Experimental Surgery and Translational Research, Biomedical Research Foundation of the Academy of Athens (BRFAA), Athens, Greece. ^52^1st Department of Neurology, Eginition Hospital, Medical School, National and Kapodistrian University of Athens, Athens, Greece. ^53^Neuromuscular/EMG service and ALS/Motor Neuron Disease Clinic, Hebrew University-Hadassah Medical Center, Jerusalem, Israel. ^54^Board of Governors Regenerative Medicine Institute, Los Angeles, CA, USA. ^55^Department of Neurology, Cedars-Sinai Medical Center, Los Angeles, CA, USA. ^56^Departments of Neuroscience, and Genetics and Genomic Sciences, Ronald M. Loeb Center for Alzheimer’s disease, Icahn School of Medicine at Mount Sinai, New York, NY, USA. ^57^Harvard Medical School, Department of Physical Medicine and Rehabilitation, Spaulding Rehabilitation Hospital, Boston, MA, USA. ^58^Center for Cellular and Molecular Therapeutics, Children’s Hospital of Philadelphia, Philadelphia, PA, USA. ^59^Department of Genetics, Perelman School of Medicine, University of Pennsylvania, Philadelphia, PA, USA. ^60^Centre for Clinical Brain Sciences, University of Edinburgh, Edinburgh, UK. ^61^Euan MacDonald Centre for Motor Neurone Disease Research, University of Edinburgh, Edinburgh, UK. ^62^Department of Genetics and Genomic Sciences, Icahn Institute of Data Science and Genomic Technology, Icahn School of Medicine at Mount Sinai, New York, NY, USA. ^63^Department of Neuropathology, Georgetown Brain Bank, Georgetown Lombardi Comprehensive Cancer Center, Georgetown University Medical Center, Washington, DC, USA. ^64^Neuroradiology Section, Department of Radiology and Biomedical Imaging, University of California, San Francisco, San Francisco, CA, USA. ^65^Neuromuscular Diseases Unit, Department of Neurology, Tel Aviv Sourasky Medical Center, Sackler Faculty of Medicine, Tel-Aviv University, Tel-Aviv, Israel. ^66^Department of Neuroscience, University of California San Diego, La Jolla, CA, USA. ^67^Department of Neurology, University of Pennsylvania Perelman School of Medicine, Philadelphia, PA, USA. ^68^Department of Neurology, Columbia University Medical Center, New York, NY, USA. ^69^Department of Pharmaceutical Chemistry, University of California San Francisco, San Francisco, CA, USA. ^70^Hadassah Hebrew University, Jerusalem, Israel. ^71^Department of Translational Neuroscience, Barrow Neurological Institute, Phoenix, AZ, USA. ^72^The Translational Genomics Research Institute (TGen), Phoenix, AZ, USA. ^73^Broad Institute, Cambridge, MA, USA. ^74^Massachusetts General Hospital, Boston, MA, USA. ^75^Institute of Molecular Medicine, Feinstein Institutes for Medical Research, Northwell Health, Manhasset, NY, USA. ^76^Korea Advanced Institute of Science and Technology (KAIST), Daejeon, South Korea. ^77^Perron Institute for Neurological and Translational Science, Nedlands, Western Australia, Australia. ^78^Integrative Immunobiology Section, National Institute of Allergy and Infectious Disease, NIH, Bethesda, MD, USA. ^79^Neuromuscular Disease Research Section, National Institute of Aging, NIH, Bethesda, MD, USA.

